# Non-invasive detection of mutagenic exposure to aristolochic acid using ultra-deep sequencing of blood and urine samples

**DOI:** 10.64898/2026.10.02.26363145

**Authors:** Peter T. Nguyen, Arnoud Boot, Po-Hung Lin, Sanskruti Sharma, Willie Yu, Mo Liu, Nanhai Jiang, Elizabeth Schmidt, Charles C. Valentine, Valerie Chew, Jesse J. Salk, Fang Yin Lo, Jacob See Tong Pang, Steven G. Rozen, Maria Zhivagui

## Abstract

Aristolochic acids (AAs) are potent nephrotoxins and carcinogens found in certain herbal remedies, driving a high burden of urothelial and liver malignancies globally. Currently, at a genomic level, AA exposure is detected retrospectively via DNA sequencing of clonal tumors, which limits opportunities to detect the mutational imprints of the exposure before tumor formation, when risk stratification may be possible. In a cohort of 12 urothelial carcinoma of the urinary tract patients, we profiled matched tumor and normal tissues alongside peripheral blood, urine sediments, and sorted leukocyte fractions. Rather than seeking tumor-derived clonal mutations, we instead interrogate non-clonal mutations associated with AA exposure. Here, we demonstrate that AA-induced mutagenesis can be non-invasively identified using ultra-deep duplex sequencing (DS), with the canonical AA signature, SBS22a, recovered in the blood and urine from patients harboring AA-positive tumors, but absent in AA-negative tumors. Single-cell WGS and DS of fractionated immune cells localized the SBS22a signal to the adaptive lymphoid compartment, identifying these long-lived cells as a systemic reservoir of mutational history. These findings show that a urothelial carcinogen leaves a durable, cell-type-resolved record in long-lived circulating lymphocytes, and establish a proof-of-concept framework for non-invasive carcinogen exposure surveillance.

## Introduction

Somatic mutations leave distinct patterns across the genome termed mutational signatures^1,2^. These signatures have traditionally been described in neoplastic tissues to delineate cancer etiologies, where clonal mutations, mutations carried by expanded clones, are readily detectable, rather than non-clonal mutations, present in individual cells or small cell populations. Although mutations in morphologically normal, non-neoplastic tissues generally remain non-clonal, recent large-scale sequencing has demonstrated that mutagenic processes are actively operational in these tissues long before clinical malignancy arises^3–5^. Characterizing these signatures in healthy cells provides a high-resolution, historical record of environmental and endogenous mutagenic exposures, offering a window for early risk stratification and population-based screening. Furthermore, the outcome of such a screen could provide tissue-specific mutational effects and inform surveillance of organs susceptible to mutagenic carcinogens^6^, drastically reducing patient burden and healthcare load^7^.

Aristolochic acids (AAs) are potent nephrotoxins and carcinogens found in certain herbal remedies, specifically plants in the genera *Aristolochia* and *Asarum*^8–11^. Exposure to AA induces a high mutation burden characterized by T>A transversions, designated as signature SBS22, driving a substantial proportion of urothelial and liver cancers globally, particularly in East Asia^12–16^. Recent studies have successfully detected SBS22 in histologically normal liver, bladder, and ureter tissues by isolating small clonal outgrowths through microdissection and characterizing their mutations using conventional DNA sequencing, capturing mutations that have undergone clonal expansion^6,17,18^. However, these approaches lack the analytical sensitivity required to resolve low-frequency mutations within highly heterogeneous, non-clonal clinical samples, such as peripheral blood or cellular sediment from urine^19–21^. To circumvent this limitation, duplex sequencing (DS), which utilizes double-stranded unique molecular identifiers to reduce PCR and sequencing artifacts to levels below one-in-ten-million based sequenced, enables the accurate detection of subclonal variants at frequencies below 0.01%^22^. A version of this technique, called bottleneck sequencing, was applied previously to detect mutagenesis associated with tobacco smoking and AA exposure in the normal kidney cortex^12^. More recently, a complementary analysis using NanoSeq, another duplex whole-genome sequencing approach, independently confirmed these findings in normal kidney samples^23^.

Here, we present a pilot study demonstrating the clinical utility of DS to non-invasively detect AA-induced mutagenesis from non-clonal mutations arising in non-neoplastic somatic cells, rather than tumor-derived mutations that have undergone clonal expansion. In a cohort of 12 patients with urothelial carcinoma arising from the bladder, ureter and renal pelvis, we show that the signature SBS22 is robustly detected in non-neoplastic tissues, peripheral blood, and exfoliated urine cells. Further investigation in peripheral blood revealed the cellular source to AA-associated mutagenesis was enriched in adaptive immune cells, rather than attributable to circulating tumor DNA. Our findings establish a direct path toward non-invasive patient screening, allowing clinicians to identify exposed cohorts, prevent ongoing toxin exposure, and initiate targeted cancer surveillance long before malignancy develops.

## Results

### Cohort overview

We collected tumor and macroscopically normal tissues from 12 urothelial cancer (UC) of the urinary tract patients. Normal tissues were collected from either tumor adjacent tissue (AN) or histologically normal tissue distant from the tumor (DN). In addition, blood (peripheral blood mononuclear cells; PBMCs) and urine samples were also collected for seven and three patients, respectively, several years after surgery (**Figure 1A**, **Supplementary Table 1**). The primary tumor sites included bladder (n=2), kidney (n=3), ureter (n=4) and kidney/ureter (n=3) (**Figure 1B**). Tumors annotated as kidney/ureter were analyzed as a combined category, as the clinical annotations were insufficient to resolve the primary site as either kidney or ureter. We profiled paired tumor and normal tissue samples using both whole-genome sequencing (WGS) and DS, whereas PBMC and urine samples were analyzed exclusively via DS (**Figure 1C**). While WGS enables broad, genome-wide mutation discovery and is well-suited for detecting clonally expanded mutations, DS was employed to detect low frequency mutations at allele frequencies below the reliable detection limit of conventional WGS, particularly in PBMC and urine, where low allele frequencies are expected. For a subset of blood samples, immune cell populations were isolated via fluorescence-activated cell sorting and subsequently characterized by single-cell WGS (scWGS) and DS, enabling high-resolution, cell-type-specific mutational profiling.

**Figure 1.**
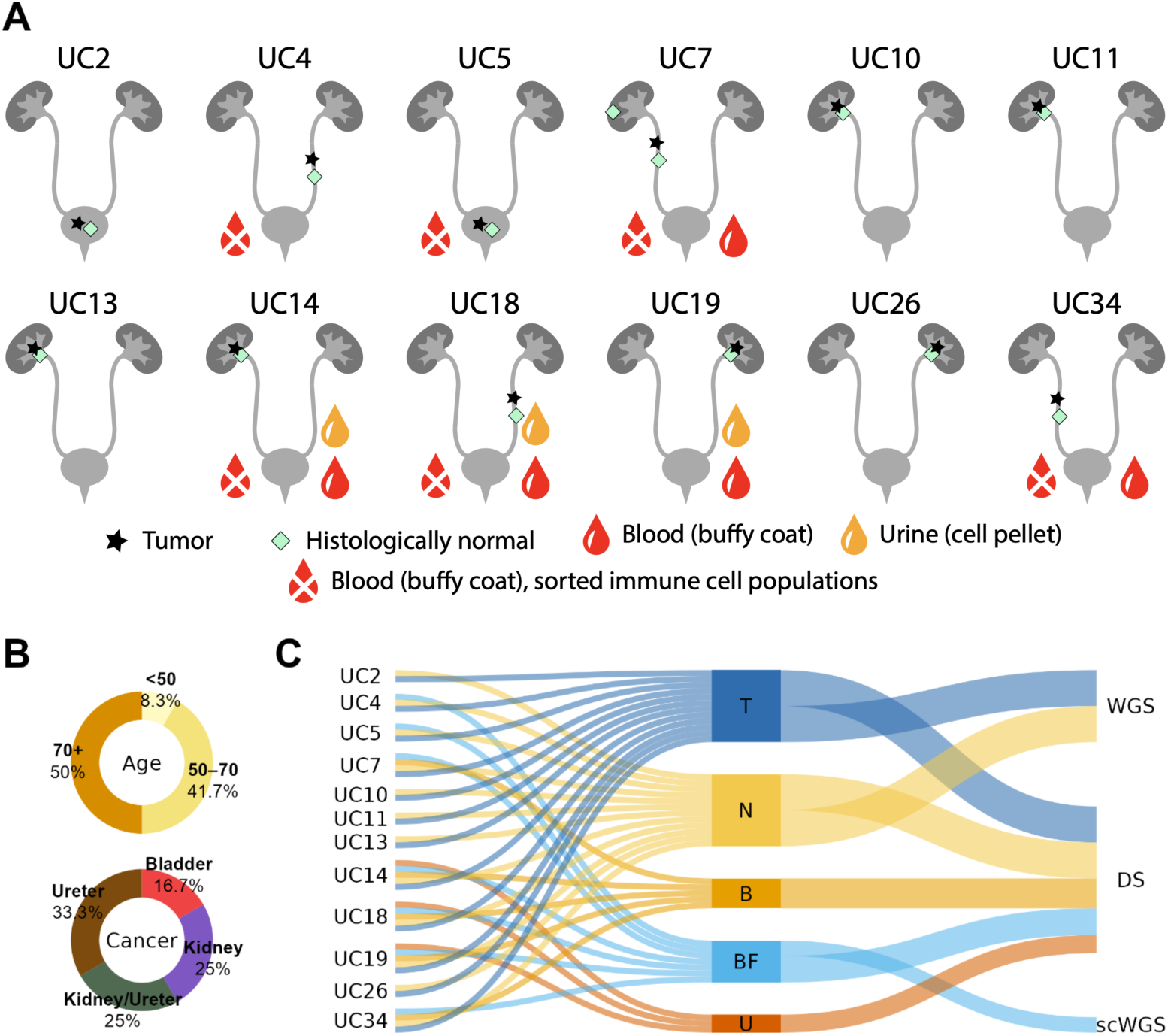
Cohort characterization. (**A**) Graphical display of samples used in this study. (**B**) Donut plot summarizing the distribution of age and cancer diagnoses. Percentages indicate the proportion of individuals in each category. (**C**) Sankey diagram illustrating the sample type and sequencing method used per patient.

### Whole-genome sequencing establishes robust SBS22a mutational signatures in primary tumor and normal tissues

Given the well-established etiological link between AA and UC^11,14,15^, we performed WGS on matched tumor-normal tissue pairs to identify patients harboring signatures of AA mutagenesis. As anticipated, mutations in the normal tissues exhibited lower variant allele frequencies (VAFs) than in the highly clonal expansions of the corresponding tumors (**Supplementary Figure 1**). In tumor samples, the median somatic mutation burden was 35,259 (IQR: 3,786–65,468) for single base substitutions (SBS), 65 (IQR: 10–252) for doublet base substitutions (DBS), and 533 (IQR: 110–811) for small insertions and deletions (IDs) (**Supplemental Table 2**). In contrast, the mutational burden in matched normal tissues was expectedly low, with a median of 110 (IQR: 67–169) SBS, 1 (IQR: 1–5) DBS, and 3 (IQR: 2–6) ID. Somatic mutation catalogues (**Supplemental Table 3-5)** were used to identify the mutational signatures through *de novo* signature extraction (**Figure 2A, Supplemental 6-8**) and attribution (**Figure 2B, Supplemental Table 9**).

**Figure 2.**
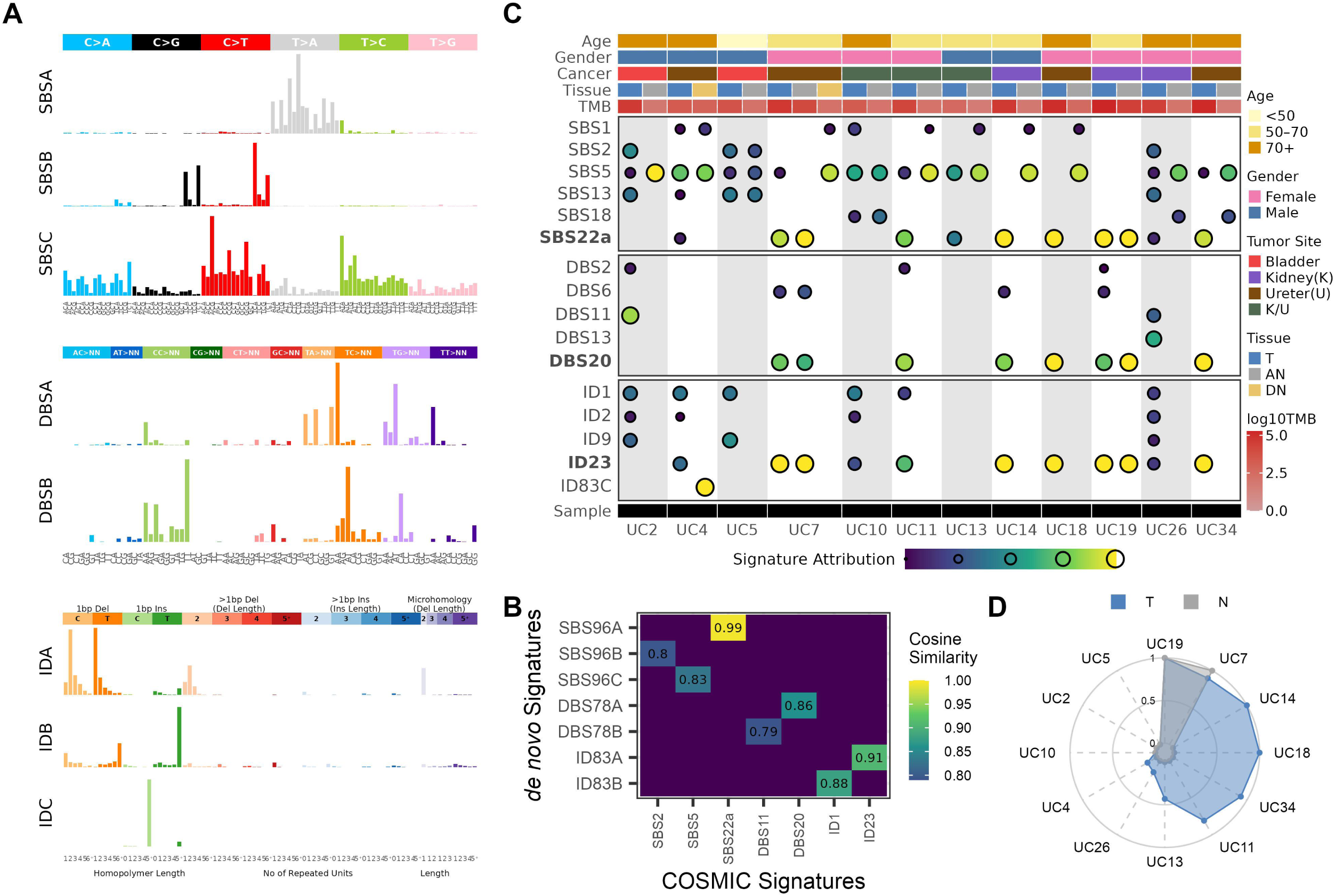
Distribution of SBS22a in whole-genome sequenced samples. (**A**) De novo signature extraction at SBS-96, DBS-78 and ID-83 level. (**B**) Heatmap showing the de novo signature extracted decomposed to COSMIC signatures with cosine similarity ≥ 0.79. Fill color indicates cosine similarity with values ≥ 0.79 displayed. (**C**) Signature attribution of COSMIC signatures, derived from *de novo* signature decomposition to mutational spectra of samples. Each point is sized and colored to represent the proportion of signature attribution in a given sample. The top annotation track corresponds to the age, gender, tumor site, tissue of origin and mutation burden for each sample. The bottom annotation track corresponds to the patient ID. (**D**) Radar plot comparing the SBS22a signature attribution between tumor and normal samples.

At the SBS level, we identified the canonical AA signature, SBS22a^24^, in nine tumors (UC4, UC7, UC11, UC13, UC14, UC18, UC19, UC26, UC34). Additionally, we observed APOBEC cytidine deaminase-associated mutagenesis, represented by signatures SBS2 and SBS13^25^ in four tumors (UC2, UC4, UC5, UC26) (**Figure 2B**-**C****, Supplemental Table 10**). Unexpectedly, SBS22a mutagenesis was also evident in the AN of UC7 and UC19, which exhibited mutational profiles highly similar to their matched tumors (cosine similarity>0.95) despite no overlap in mutations. This lack of shared variants suggests that the AA-associated mutational processes operated independently in these distinct tissue compartments rather than arising from a common progenitor clone. Consistent with the repair of bulky dA-aristolactam adducts on adenines^12,13^, we observed a pronounced transcriptional strand bias for T>A substitutions in both the tumor and normal tissues harboring SBS22a, with a significant enrichment of mutations occurring on the transcribed strand (**Supplementary Figure 2, Supplemental Table 11**).

We next evaluated DBS and ID to determine whether these orthogonal mutational features corroborate with the SBS findings, thereby providing an integrated view of the mutational landscape. We restricted mutational signature attribution to samples with at least 25 mutations, consistent with the low-mutation sample filtering implemented in mSigAct. The AA-associated DBS signature, DBS20^24^, was detected in nearly all SBS22a-positive samples. Likewise, we observed high concordance between the SBS findings and the AA-associated ID signature ID23^24^ (**Supplemental Figure 3, Supplemental Table 12**). Crucially, neither DBS20 nor ID23 was attributed to the SBS22a-negative samples (UC2 and UC5), demonstrating the high specificity of these multi-class genomic scars and further verifying AA exposure in SBS22a-positive cases.

Together, comprehensive mutational signature profiling of the WGS data defined the cohort’s AA exposure status, establishing a high confidence reference set for all subsequent liquid biopsy analyses.

### Ultra-deep duplex sequencing uncovers persistent SBS22a signature dominantly in non-neoplastic tissues

A limitation of WGS is its reduced sensitivity for low-input samples like PBMC and urine, where mutant alleles are expected to be present at ultra-low frequencies. Utilizing our WGS-characterized cohort, we investigated whether the AA-associated mutational signatures could be recapitulated using DS, thereby validating the robustness of the AA mutational signature and confirming its detectability using an orthogonal, ultra-sensitive sequencing platform. We performed DS on all WGS-profiled samples, as well as matched PBMC and urine samples where available using TwinStrands Human Mutagenesis panel, which is strategically designed to survey genomic regions that broadly reflects the trinucleotide context distribution of the whole genome while systemically avoiding confounding hotspots and non-randomly mutated genomic regions^22,26^ (**Figure 1A**). The median tumor mutational burden with DS was 218 (IQR: 187–428) for SBS, 2.5 (IQR: 1–5.5) for DBS, and 22.5 (IQR: 12.5–37.8) for ID (**Supplemental Table 13**). Intriguingly, and in contrast to our WGS observations, the SBS and DBS burdens in normal tissues were highly comparable to those of their matched tumors, with a median of 238 (IQR: 210–309) SBS (Wilcoxon rank-sum test, *p*=0.66) and 3 (IQR: 1–4.3) DBS (*p*=1.00). However, the median normal tissue ID burden (9.5; IQR: 8–13.3) remained significantly lower than that observed in the tumors (*p*=0.03).

Due to the limited number of samples exceeding the 25-mutation threshold implemented in mSigAct for DBSs and IDs, specifically 2 and 8 out of 32 samples, respectively, these mutation classes were not analyzed (**Supplemental Figure 4-5, Supplemental Table 14-16**). At the SBS level, we detected SBS22a in all normal tissues matched to patients whose tumors exhibited SBS22a by WGS (Fisher’s exact test, *p*=0.045) (**Figure 3A-B, Supplemental Table 17**). Intriguingly, the majority of the tumors (9 out of 12) did not exhibit detectable SBS22a by DS. This is highly consistent with clonal evolution: as a tumor grows, a single clone expands and dominates the tissue, rendering the tumor genetically homogeneous. This uniformity reduces the diversity of unique, low-frequency mutations captured by DS, whereas the normal tissue remains a highly diverse mosaic of independent cells that preserve the AA signature.

**Figure 3.**
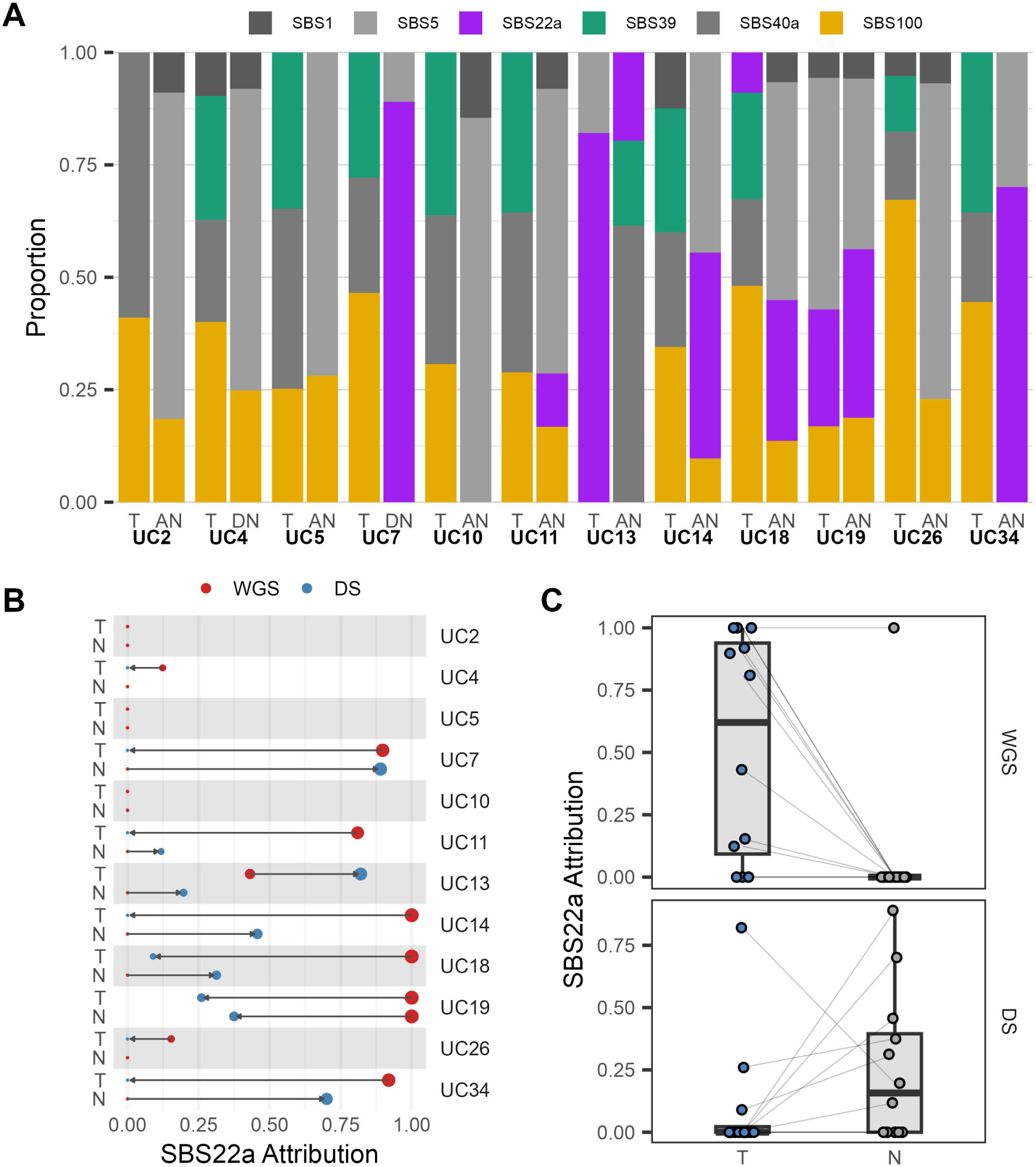
Distribution of SBS22a in duplex sequenced samples. (**A**) Signature attribution of COSMIC signatures, derived from *de novo* signature decomposition to mutational spectra of samples. Y-axis represents the relative proportion of each signature to the mutational spectra for each sample, with the color bar corresponding to the individual mutational signature. Bars are grouped by patient and stratified by sample type. (**B**) Dumbbell arrow plot comparing SBS22a attribution between WGS and DS, grouped by patient and stratified by sample type. Each point is colored by sequencing method and sized by SBS22a attribution level. Arrow denotes the directional change from WGS to DS. Arrow length reflects the magnitude change. (**C**) Boxplot summarizing SBS22a attribution between tumor and normal samples by sequencing method. Each point represents a single sample, while connected to its paired sample.

Notably, the tumor of UC13 was the sole exception, displaying a prominent SBS22a signature by DS despite demonstrating minimal SBS22a mutagenesis by WGS (**Figure 3B-C, Supplemental Table 10, Supplemental Table 17**). Furthermore, in this patient, DS revealed a higher SBS22a burden in the tumor relative to the AN tissue, with 1,161 SBS22a-associated mutations detected in the tumor compared to 48 in the normal. This profile suggests a late-stage exposure to AA during tumor development, allowing a high volume of subclonal, non-clonal SBS22a-associated mutations to accumulate in the tumor^27^. In contrast, the lower mutational density in the matched normal tissue likely reflects slower cellular turnover, which limits the replication-dependent fixation of DNA adducts into detectable somatic mutations^28^.

SBS39^2^ was enriched in tumors, occurring in 75% of the tumor samples, but in only one normal sample (Fisher’s exact test, *p*=0.0014) (**Figure 3A**). Although its etiology remains unknown in COSMIC, a recent study proposed an association with homologous recombination deficiency (HRD)^29^. However, we detected no corresponding canonical HRD-associated signatures in our WGS data^2,25,30^, suggesting that its enrichment is independent of HRD pathways. Additionally, SBS100^31,32^, a predominantly C>A signature, was detected in the majority of both tumor and normal samples (79%, 19 out of 24; exact binomial test, *p*=0.0033). Given its widespread presence, SBS100 likely reflects a background process or technical artifact, such as C>A oxidative DNA damage introduced during sample preparation^33^, rather than *bona fide* tobacco-induced mutagenesis.

Collectively, DS reliably recapitulated individuals with strong SBS22a attribution by WGS, with SBS22a detected in normal tissues of SBS22a-positive tumors by WGS. Notably, SBS22a was not detected by DS in two patients, despite exhibiting SBS22a in the tumor by WGS. In UC4, this may be attributable to spatial heterogeneity, as DN tissue may not have experienced the same local AA-associated mutagenic assault as the tumor environment, whereas UC26 exhibited a weaker AA-associated signal in the tumor by WGS (15.3%), reducing the likelihood of detection by DS.

### Duplex sequencing enables non-invasive detection of SBS22a in liquid biopsies

We next investigated whether DS could detect AA-associated mutagenesis in liquid biopsies, thereby enabling opportunities for early risk stratification and population-based screening. Given that our cohort consisted of patients with UC of the urinary tract, we examined urine samples as a biologically relevant source to determine whether epithelial cell shedding into the urine, which has gained increasing interest for non-invasive cancer detection^34,35^, could capture the AA-associated mutagenesis observed in neoplastic and non-neoplastic tissues. To test this, we performed DS on three urine sediments from UC14, UC18, and UC19 (**Figure 4A**), and observed a median SBS mutation burden of 148 (IQR: 81–214) (**Supplemental Table 13**). The remaining samples could not be analyzed because they were not collected from those individuals. Using the same signature attribution approach in our tumor and normal samples, we detected SBS22a in the urine of these patients, all of which harbored SBS22a-positive tumors by WGS (**Figure 4B, Supplemental Table 16-17**). Restricting the analysis to T>A mutational profile, UC19 showed the highest similarity to the de novo signature (cosine similarity=0.95) (**Figure 4C**), whereas the other samples had fewer mutations and consequently less well-resolved SBS22a mutagenesis. We next compared SBS22a attribution levels between matched urine and tissue samples. We observed a strong association with non-neoplastic tissue (R^2^=0.98), whereas neoplastic tissue was weaker (R^2^=0.73; n=3) (**Figure 4D-E**).

**Figure 4.**
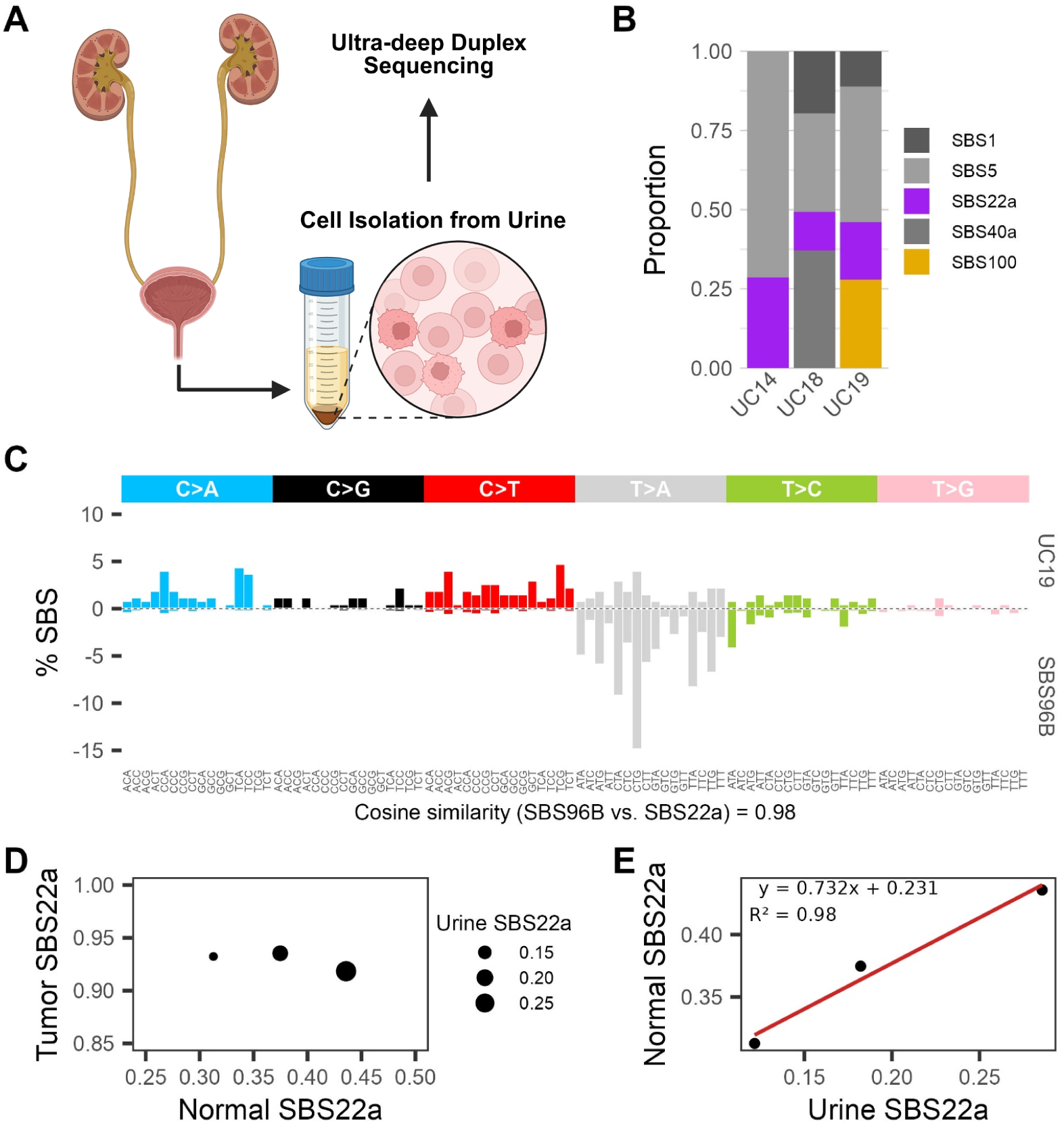
Detection of SBS22a in urine. **(A)** Schematic illustration for isolating cell populations from urine prior to duplex sequencing. **(B)** Signature attribution of COSMIC signatures, derived from *de novo* signature decomposition to mutational spectra of samples. Y-axis represents the relative proportion of each signature to the mutational spectra for each sample, with the color bar corresponding to the individual mutational signature. Bars are grouped by patient and stratified by sample type. **(C)** SBS-96 mutational signature plot comparing UC19’s urine mutational spectrum with SBS96B (*de novo* signature). Cosine similarity scores reflect similarity between SBS96B and SBS22a. **(D)** Scatter plot comparing SBS22a attribution between tumor whole-genome sequenced and normal duplex sequenced. Each dot represents a single sample and is sized according to their SBS22a attribution in the urine. **(E)** Scatter plot comparing SBS22a attribution between urine and normal duplex sequenced. Fitted linear regression line with equation and R^2^ value are shown.

Having established that urine can capture AA-associated mutagenesis, we next determined whether PBMC, a systematic source of circulating cellular components, recapitulates the same signature attribution. To test this, we performed DS on five PBMCs (UC7, UC14, UC18, UC19, UC34) and observed a median SBS mutational burden was 303 (IQR: 279–307) (**Supplemental Table 13**). The remaining samples could not be analyzed because they were not collected from those individuals. Given that the liquid biopsies were collected without prior knowledge to AA status, our study did not include samples from SBS22a-negative tumors by WGS. We therefore included an independent dataset containing negative samples generated using NanoSeq^23^ with our analysis to further evaluate the specificity of SBS22a detection (**Figure 5A-B**). We reanalyzed these samples using our signature attribution approach, and identified SBS22a-positive (n=47) and negative samples (n=18) (**Figure 5A**). Similar to our urine analysis, we successfully detected SBS22a in all blood samples, all of which harbored SBS22a-positive tumors by WGS, in our study (**Figure 4A, Supplemental Figure 6, Supplemental Table 17**). We also observed a similar trend with the independent NanoSeq dataset. We observed SBS22a in PBMC of SBS22a-positive samples, although not observed in all samples (44.7%). In contrast, only 3 out of 18 SBS22a-negative samples (16.7%) showed SBS22a attribution. When comparing the SBS22a attribution of positive and negative samples, we observed a significant difference (Wilcoxon rank-sum test, *p*=0.03), establishing that SBS22a attribution separates AA-exposed from unexposed blood while underscoring that reliable detection requires ultra-deep coverage. The lower detection rate in these samples is likely attributable to their considerably lower sequencing depth (∼20x NanoSeq vs ∼6,300x this study), reducing opportunities to detect low-frequency mutations. Based on the empirically derived median VAF of T>A mutations (0.00989%) across the five PBMC samples in this study, Poisson sampling estimated the probability of observing at least one mutation is 0.20% at 20x coverage, compared to 46.4% at 6,300x. We observed no significant correlation between SBS22a levels in matched blood and tissue samples (R^2^>0.26). This quantitative divergence likely reflects tissue-specific differences in AA pharmacokinetics, cell turnover rates, and clonal dynamics between the hematopoietic system and the urological epithelium. Nevertheless, the reliable detection of SBS22a in blood and urine demonstrates the technical feasibility of identifying historical AA exposure using minimally invasive clinical specimens.

**Figure 5.**
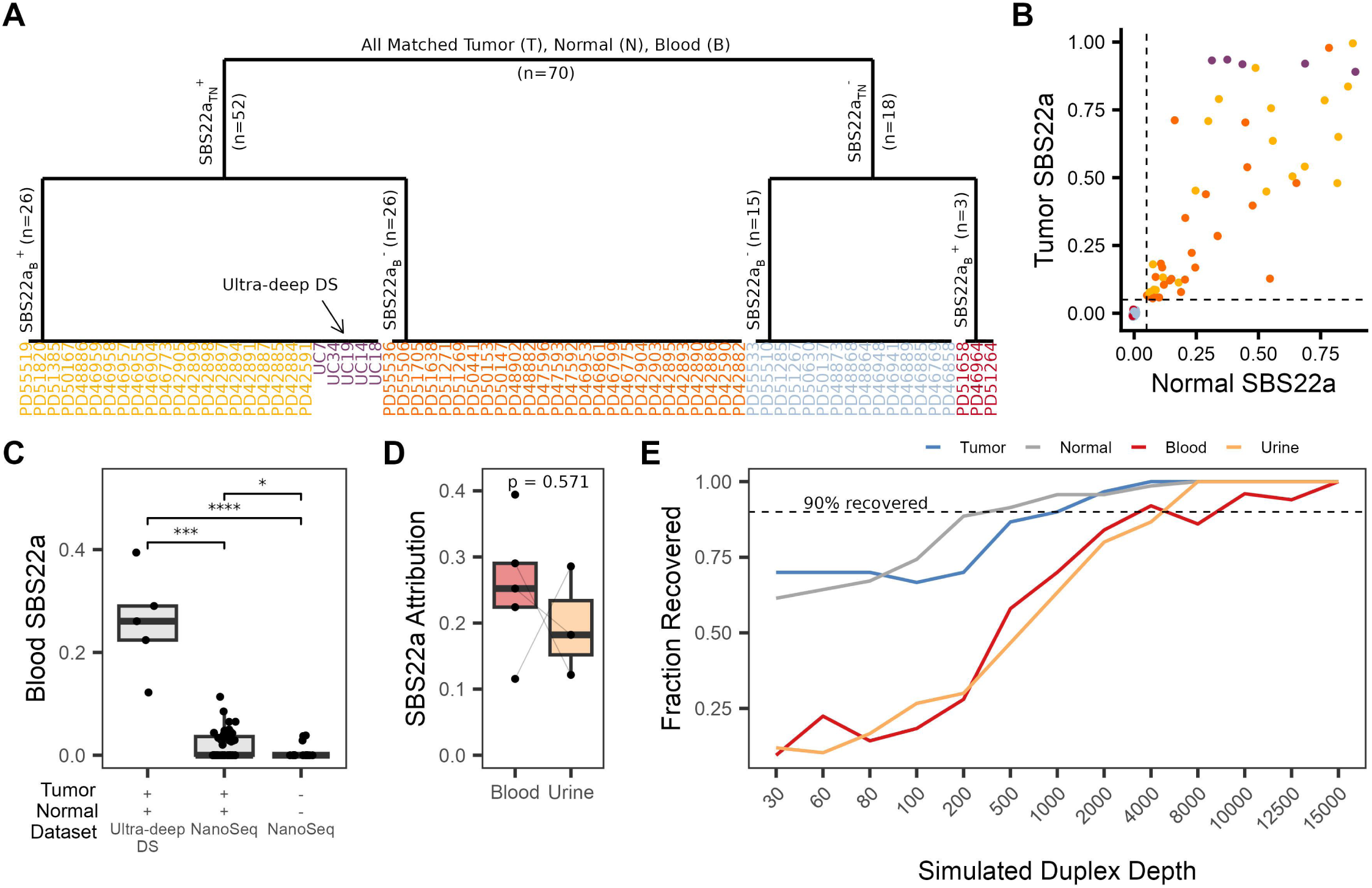
Detection of SBS22a in blood. **(A)** Dendrogram illustrating the hierarchical clustering based on SBS22a presence in tumor, normal and blood samples by duplex sequencing or NanoSeq. **(B)** Scatter plot comparing SBS22a attribution between tumor whole-genome sequenced and normal duplex sequenced. Each dot represents a single sample. Dashed horizontal and vertical lines indicate the 5% threshold used to define signature presence. **(C)** Boxplot summarizing SBS22a attribution in blood. Each point represents a single sample, stratified by sequencing method and AA status determined by signature attribution with significance levels using the Wilcoxon rank sum test indicated by asterisks (*, p<0.05; **, p<0.01; ***, p<0.001; ****, p<0.0001). **(D)** Boxplot summarizing SBS22a attribution in blood and urine duplex sequenced. Each point represents a single sample, while connected to its paired sample. Statistical significance was calculated using the Wilcoxon rank sum test. **(E)** Trace plot summarizing the duplex depth simulation analysis to determine the minimum duplex depth required for robust SBS22a detection across sample type. Black dashed line represents the minimum duplex depth to recover SBS22a in 90% of resampled SBS22a-positive samples.

Moreover, we evaluated whether robust signature detection could be maintained at lower sequencing depths to optimize clinical utility and cost-efficiency for screening. To address this, we performed an *in silico* downsampling simulation across 13 target coverage tiers, ranging from 30x to 15,000x (**Figure 4B, Supplemental Table 18**), modeling alternative duplex consensus read counts based on the observed VAFs of each sample. While SBS22a remained detectable in tumor and normal tissues at low sequencing depths, PBMC and urine samples required substantially higher coverage due to their low mutant allele fractions, requiring approximately 4,000x and 8,000x duplex depth, respectively, to recover SBS22a in 90% of resampled SBS22a-positive samples. Beyond these thresholds, signature recovery rates plateaued, indicating a clear limit of diminishing analytical returns relative to increased sequencing costs. Together, these findings demonstrate that the optimal sequencing depth for exposure detection is highly compartment-specific, a critical design parameter for future non-invasive biomarker assays.

### Clonal enrichment of SBS22a in solid tissues versus polyclonal biofluids

Recent studies have demonstrated that histologically normal human urothelium is colonized by macroscopic clonal expansions^18,36^, which explains our ability to detect SBS22a in normal tissue WGS profiles. To characterize these subclonal dynamics in our DS data, we evaluated whether we could identify signatures of clonal expansion within the normal urothelial samples. For this analysis, we restricted our somatic call set to variants supported by at least two independent duplex consensus reads, hereafter operationally defined as clonal mutations, reflecting the presence of mutation in multiple independent DNA molecules consistent with clonal expansion. Based on hypergeometric sampling using the average DS sequencing depth, we calculated that any somatic clone constituting at least one-in-one-thousand (0.1%) of the cells within the biopsy sample would be robustly detected by at least two reads at our target sequencing depth, with the probability plateauing at approximately 1 as the proportion of mutant cells increases (**Supplemental Figure 7, Supplemental Table 19**). Utilizing this filtered call set, we performed mutational signature attribution to resolve the active mutational processes driving these subclonal expansions in normal tissues.

We observed a strong negative correlation between the proportion of clonal SBS22a mutations and the expected gradient of cellular heterogeneity, increasing from tumor to normal, blood and urine (Spearman’s rho=-0.80, *p*=5.82×10^-5^) (**Figure 6A, Supplemental Table 20**). This is consistent with tumor clonal expansion propagating SBS22a-associated mutagenesis across a larger fraction of cells. Furthermore, the presence of non-clonal SBS22a mutations across all sample types demonstrates that AA-induced mutations occur broadly across cells that are too infrequent to be detected by conventional WGS. Conversely, the proportion of clonal, aging-associated mutations showed a weak non-significant positive correlation with expected heterogeneity (Spearman’s rho=0.36, *p*=0.075) (**Figure 6B**). Furthermore, SBS22a levels was significantly enriched in clonal mutations compared to non-clonal, occurring in 92% of tissue specimens, whereas PBMC and urine samples remained highly polyclonal as indicated by the negligible clonal enrichment (25%) (**Figure 6A**).

**Figure 6.**
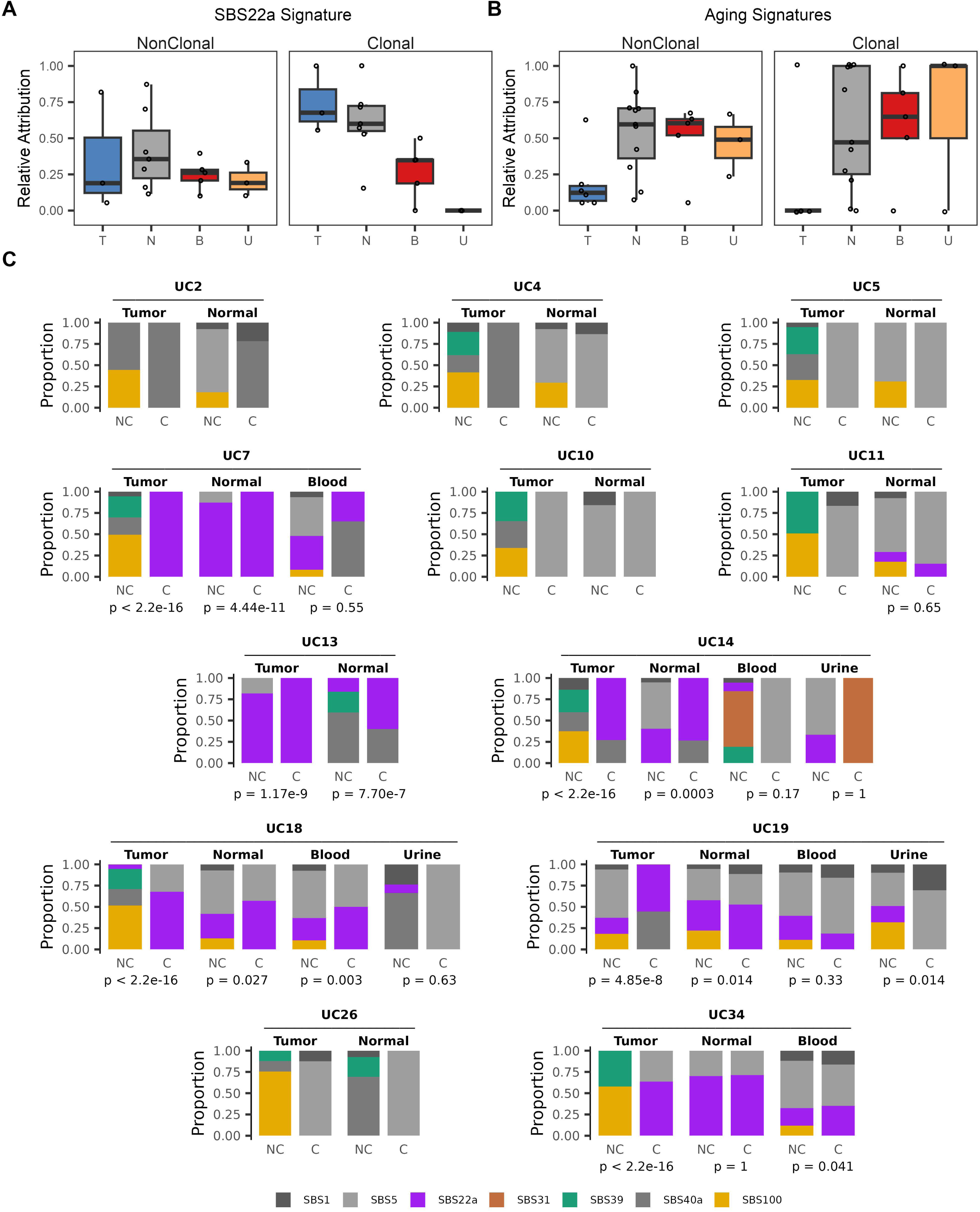
SBS22a mutagenesis contributes to clonal expansion. (**A**) Boxplot comparing the proportion of SBS22a and (**B**) aging-associated mutations (SBS1+SBS5) that are supported by 1 read (NonClonal; NC) and at least 2 reads in the DS data (Clonal;C) stratified by the expected gradient of cellular heterogeneity, increasing from tumor to normal, blood and urine. (**C**) Signature attribution of COSMIC signatures, derived from *de novo* signature decomposition to mutational spectra of DS samples using the nonclonal (“NC”, left) and clonal mutations (“C”, right). Enrichment of SBS22a in clonal mutations was performed using a binomial test, where p is the proportion of SBS22a mutations in the nonclonal DS spectrum (“NC”). P-values are plotted below the plot.

### Systemic sharing of somatic variants between blood and tissue specimens

Given AA presence in PBMCs, we next investigated the cellular provenance, as signals could have potentially originated from circulating tumor DNA (ctDNA). We crossed somatic variants in PBMCs against those detected in the corresponding tumor by WGS and observed no overlap, suggesting the mutations are not tumor-derived. However, we observed partial overlap between variants detected in blood and the remaining patient-specific samples (**Supplemental Figure 8**), suggesting these variants are native to the hematopoietic system, and possibly reflect leukocyte infiltration. If this holds true, variants shared across compartments should be enriched for true clonal events within the blood. Consistently, while only 11.4% of all blood variants met our clonal threshold, 45.4% of the variants detected across the blood and other sample types were clonal in the blood, supporting the hypothesis that AA mutagenesis occurs natively within PBMCs, acting as a benign clonal recorder of historical exposure. This suggests that peripheral blood can be leveraged as a systemic sentinel compartment to reconstruct lifetime toxicological exposures, offering a powerful alternative to invasive tissue biopsies.

### SBS22a accumulation is enriched in lymphoid cell populations in blood

To investigate the cellular source of AA mutagenesis in PBMCs, which must arise from circulating nucleated cells if not ctDNA, we fractionated PBMCs into B-cell, T-cell, NK-cell and myeloid subpopulations and performed scWGS (**Figure 7A**). In total, we whole-genome sequenced 27 single cells isolated from six different patients, comprising of five individuals with SBS22a-positive tumors (UC4, UC7, UC14, UC18, UC34) and one SBS22a-negative control (UC5). The remaining samples could not be analyzed because they were not collected from those individuals. To mitigate single-cell sequencing artifacts, we applied *SigRescueR* using six untreated scWGS cell lines for background correction as their mutational patterns are expected to predominately reflect technical artifacts introduced by scWGS (**Supplemental Figure 9, Supplemental Table 21**). Computational filtering classified an average of 74% of raw somatic calls as technical background (**Supplemental Figure 9B**). Following artifact removal, the deconvoluted mutational spectra exhibited a highly resolved SBS22a signal relative to the uncorrected profiles (**Supplemental Figure 9A**, **Supplemental Figure 10, Supplemental Table 22-23**). Subsequent *de novo* signature extraction and attribution confirmed the presence of SBS22a in 12 single cells (**Figure 7B, Supplemental Table 24-25**). Next, we assessed whether variations in sample processing times introduced batch effects. We observed a nominal enrichment for SBS36 and SBS45 (OR=18.8 and 6.4, respectively) in one batch, which did not remain statistically significant after multiple-hypothesis testing correction. Genomic analysis revealed no evidence of deleterious *MUTYH* variants or deficiency in these samples. Furthermore, using D-ToxoG, we confirmed that these C>A transitions lacked the characteristic read-strand bias indicative of the SBS45 artifact^33^. We therefore conclude that this signature represents a spurious, batch-specific artifact rather than reflecting true endogenous biology.

**Figure 7.**
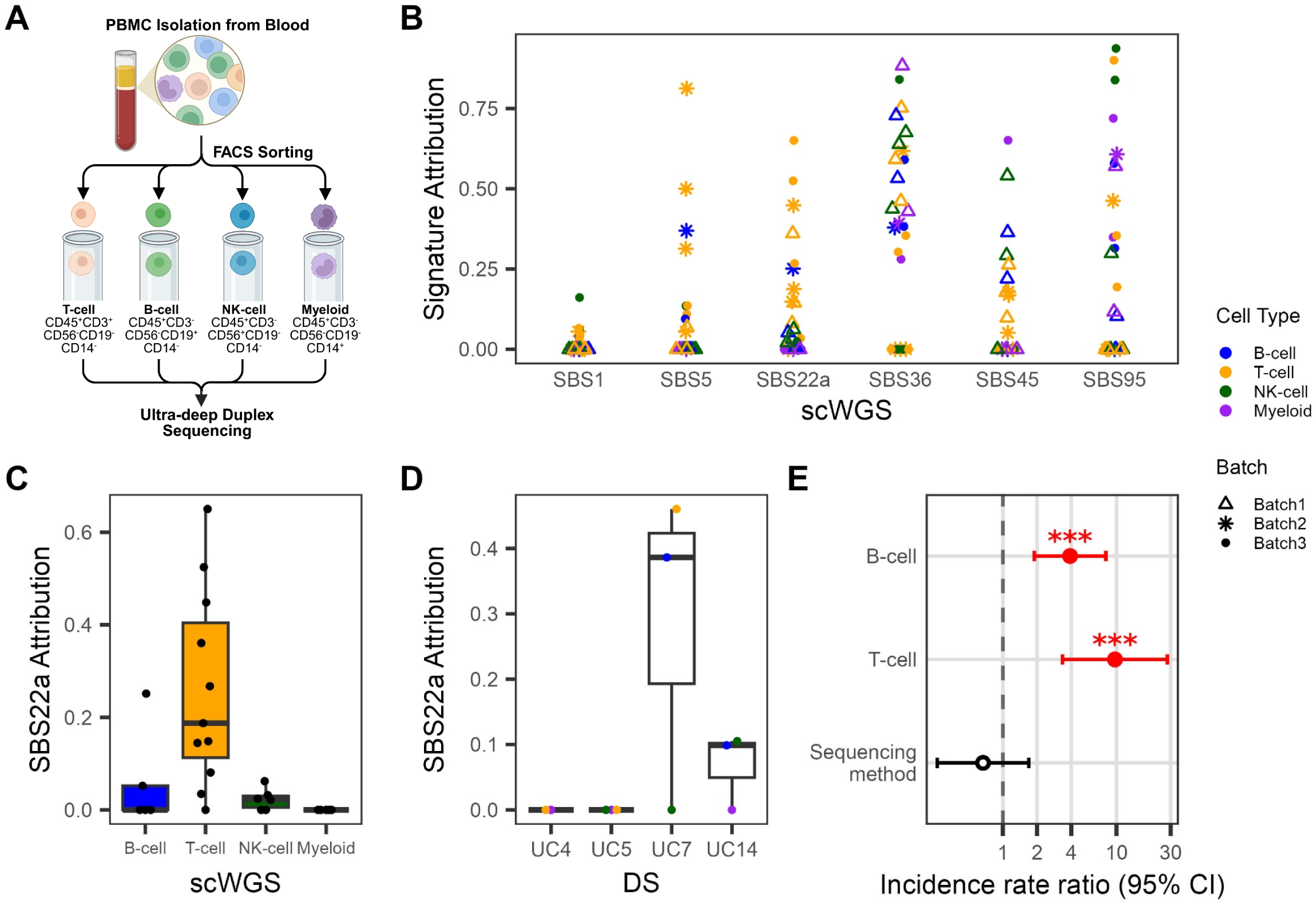
SBS22a is enriched for lymphatic cells. **(A)** Schematic illustration for isolating and sorting immune cell populations from PBMCs prior to duplex sequencing. **(B)** Signature attribution of COSMIC signatures, derived from *de novo* signature decomposition to mutational spectra of patient-derived immune cells scWGS. Each point represents a single-cell immune cell, with shape indicating sequencing batch and color denoting immune cell type. **(C)** Boxplot summarizing SBS22a attribution patient-derived immune cells scWGS. Each point represents a single-cell immune cell, stratified by immune cell population. **(D)** Boxplot summarizing SBS22a attribution of duplex sequenced sorted immune cell samples. Each point represents a single sample, stratified by patient and colored by immune cell type. **(E)** Forest plot showing adjusted incidence rate ratios (IRRs) and 95% confidence intervals for SBS22a burden relative to NK-cells (reference group). The GLM model included immune cell type, sequencing platform, and total mutational burden as covariates. Cluster-robust standard errors were used to account for patient-level clustering. The vertical dashed line indicates an IRR of 1 (no difference relative to the reference group). Red points denote statistically significant associations (p<0.05), with significance levels indicated by asterisks (*, p<0.05; **, p<0.01; ***, p<0.001).

To complement our single-cell findings and capture low-frequency mutations that may have escaped scWGS sampling, we performed DS on bulk-sorted immune cell populations from four patients (UC4, UC5, UC7, UC14). The remaining samples could not be analyzed because they were not collected from those individuals. Prior to signature attribution, we applied *SigRescueR* to remove SBS31^37^, a platinum-based chemotherapy-associated signature, from immune cell fractions originating from UC14 due to the patient receiving platinum-based chemotherapy as part of their standard-of-care treatment (**Supplemental Figure 11, Supplemental Table 26-28**). For UC7, DS confirmed the presence of SBS22a in B-cells and T-cells, but absent in NK-cells (**Figure 7C, Supplemental Figure 12, Supplemental Table 29**). For UC14, SBS22a was robustly detected in B-cells and additionally captured within the NK-cell fraction. Crucially, none of the sorted cell fractions from UC5 exhibited detectable SBS22a mutagenesis, confirming the high specificity of our detection pipeline (**Figure 7D**). Intriguingly, SBS22a was not captured by DS in UC4. Given that this individual displayed both low signature attribution in the primary tumor by WGS and only borderline signal in T-cells via scWGS, this suggests that the underlying AA-induced mutational process in this patient fell below detection threshold.

To evaluate immune cell-specific SBS22a accumulation, we fitted a negative binomial generalized linear model (GLM) across immune cell populations while controlling for total mutation burden and sequencing technology (**Figure 7, Supplemental Table 30**). Myeloid cells were excluded from the GLM analysis due to a lack of SBS22a mutations, resulting in an unestimable boundary effect. Relative to NK-cells, SBS22a burden was significantly enriched in B-cells (IRR=3.90; CI: 1.80-8.06; *p*<0.001) and T-cells (IRR=9.69; CI:3.34-28.06; *p*<0.001). Sequencing platform was not significantly associated with SBS22a burden (IRR=0.67; CI: 0.26-1.69; *p*=0.39), suggesting the difference was not driven by platform-specific effects. Despite the small number of patient clusters (n=6), our findings suggest the mutational processes driving SBS22a activity are highly cell-type specific, preferentially detectable in the adaptive lymphoid compartment (T- and B-cells), most consistently in T-cells, and below the detection threshold in innate myeloid populations.

### High-depth validation of lineage-specific somatic variants in sorted blood cells

To assess the clonal architecture of these mutations, we compared the variants identified in the bulk-sorted leukocyte fractions of UC7 and UC14 to those detected in the corresponding unsorted bulk PBMCs (**Supplemental Table 31**). Variants recaptured across both DS assays were strongly enriched for mutations classified as ‘clonal’ in the initial bulk experiment (OR=15.7 for UC7, OR=27.9 for UC14; *p*=2.24×10^−16^ and 1.39×10^−12^, respectively, Fisher’s exact test; **Supplementary Figure 13**). Notably, the vast majority of somatic variants were strictly restricted to a single leukocyte lineage. In patient UC7, 70.4% of variants were captured exclusively in T-cells, whereas in patient UC14, 82.1% of variants were unique to NK-cells. Accordingly, only 7.95% (UC7) and 11.4% (UC14) of variants were shared between different cell fractions from the same individual. This striking lineage isolation is consistent with somatic mutagenesis occurring during localized clonal expansions in circulation, likely driven by immune activation. Crucially, because none of the sparse mutations shared between distinct cell fractions exhibited the characteristic T>A transversion, we conclude that the SBS22a footprint does not originate from early, multi-lineage hematopoietic progenitors.

### A model of AA exposure history based on sequencing data

Altogether, we used WGS and DS to characterize AA exposure in tumor, normal, blood and urine samples by interrogating clonal and non-clonal mutations, respectively (Figure 8A). The combination of these complementary sequencing approaches provides support for two models of exposure history, consistent with differences in timing and persistence (Figure 8B). The first model represents chronic exposure to AA before tumor development. In this model, individuals are unknowingly exposed to AA, resulting in mutagenesis prior to cancer initiation. These AA-associated mutations are clonally expanded and detectable in both tumor and normal tissues by WGS. Continued exposure to AA during tumor development would introduce additional mutations, contributing to the WGS profile, while also generating rare, non-clonal mutations detected by DS. In the second model, AA exposure occurs after tumor initiation, accumulating in the tumor and contributing to the WGS profile, while also generating rare, non-clonal mutations detected by DS. Furthermore, the presence of AA-associated mutagenesis in DS may indicate recent or current exposure, as the absence from DS may reflect prior exposure that have clonally expanded reducing detection by DS. Discussion

**Figure 8.**
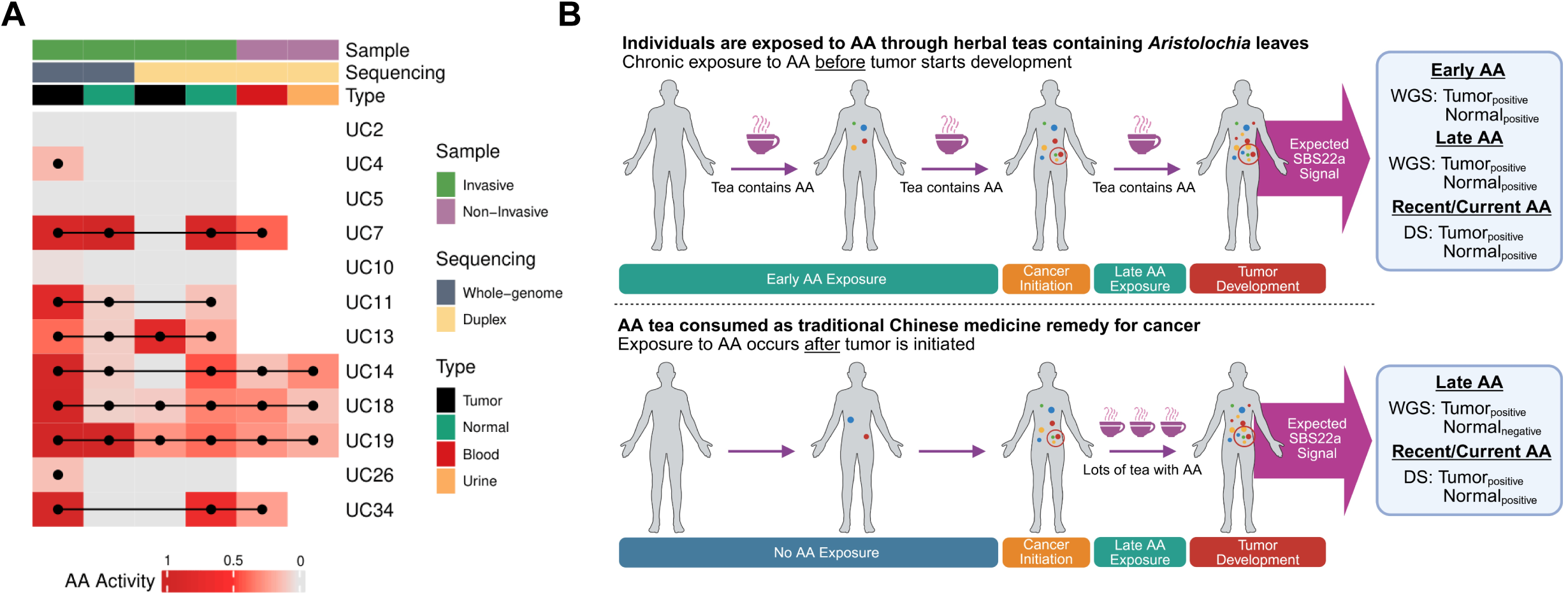
Proposed Model of AA Exposure based on WGS and DS. (A) Heatmap showing the relative signature attribution of COSMIC signatures to mutational spectra of samples. Each dot represents a sample with positive SBS22a attribution, with lines connecting samples from the same patient. The top annotation track corresponds to specimen type, sequencing method and sample type. (B) Proposed AA exposure history model.

AA induces a distinctive mutational signature, enriched by T>A transversion^8–16^. Although early detection strategies, such as urine-based assays, have emerged as a non-invasive alternative to tissue biopsies, they primarily target tumor-specific genes rather than the underlying mutational processes that precede it^38^. We sought to determine whether the systemic footprint of AA-associated mutagenesis could be non-invasively intercepted using blood or urine, establishing a framework for pre-clinical carcinogen exposure screening.

By profiling matched tumor, normal, blood, and urine samples from 12 UC patients, we observed SBS22a detection that varied across sequencing methods. Tumors that were SBS22a-positive by WGS were absent by DS; conversely, normal tissues that were SBS22a-positive by DS went undetected by WGS^19,22^. The observed differences in SBS22a levels between WGS and DS are likely attributable to sequencing design and tissue architecture^21,39^, with WGS optimized to capture high-frequency clonal somatic variants, making it ideal for tumors, whereas DS enabled detection of low-frequency mutations in normal tissues which undergo limited cell proliferation. Additionally, in duplex sequenced tumors, we observed two distinct scenarios, the absence or presence of SBS22a, potentially reflecting prior vs ongoing exposure states, respectively.

In UC7, conventional WGS resolved a strong SBS22a signal exclusively within the AN tissue, while the DN tissue remained negative. While recent studies have circumvented the lack of clonality in normal tissues by taking small patches of cells to enrich for single clones^40,41^, this approach is not possible for blood and urine, reinforcing the necessity of duplex architectures for non-invasive detection.

Using DS, we robustly detected SBS22a in all matched blood and urine samples from patients with AA-positive tumors by WGS, supporting the feasibility of a non-invasive screen for mutagenic carcinogens. Furthermore, the overlap of somatic variants shared between tissue samples and blood collected years apart reinforces the hypothesis of long-lived AA-mutagenized lymphocytes contributing to blood production in circulation, highlighting the utility of blood for non-invasive screening^42^.

The enrichment of SBS22a in lymphoid cells raises the question why these cells preferentially accumulate AA-induced mutagenesis. One mechanism consistent with this pattern is the fixation of persistent dA-aristolactam adducts on adenines during lymphocyte proliferation. AA is systemically bioavailable and forms aristolactam adducts in non-target organs, including bone marrow^43,44^. The 7-(deoxadenosin-N6-yl)-aristolactam I adduct evades global-genome nucleotide excision repair and persists in humans for over two decades after exposure ceases^45–47^. Conversion of this stable adenine adduct into a fixed T>A substitution requires DNA replication. Among circulating leukocytes, adaptive lymphocytes undergo the most extensive antigen-driven proliferation. Single-cell sequencing shows that memory B- and T-cells carry higher mutation burdens than naive and hematopoietic stem cells, with T-cells accumulating mutations at the highest lifelong rate^48,49^. This proliferative history is consistent with the observed B- and T-cell enrichment over NK-cells. The absence of SBS22a in myeloid cells is explained by their short circulating half-life of 1-2 days and continuous replacement from the stem-cell pool, which prevents retention of adduct-derived mutations, whereas long-lived memory lymphocytes retain them^50,51^.

We note several limitations to our work. First, patient-level AA exposure history was unavailable, reflecting a broader challenge in exposure assessment, as individuals are rarely aware of prior exposure. We addressed this by inferring exposure directly from the presence of highly specific computational mutational signatures (SBS22a, DBS20, and ID23) by WGS. Second, our cohort was constrained to 12 patients, with matched blood and urine available for seven and three individuals, respectively. Although these samples provide a unique matching datasets, the limited sample size can lead to unstable variance estimates, potentially affecting the precision of the standard errors, confidence interval and p-values, and thereby, reducing statistical power, highlighting the need for larger cohorts to validate these associations. Finally, matched blood and urine samples were only available from individuals whose primary tumors were AA-positive. The absence of an AA-negative cohort with matched liquid biopsies limits our ability to formally define the analytical specificity of our screening approach. We attempted to address this by analyzing an independent dataset with matched tumor, normal and blood samples with AA-negative and positive samples^23^, and although not all AA-positive samples had SBS22a attribution, we observed differences in SBS22a levels between positive and negative samples. This observation might be attributable to a considerably lower depth in the independent dataset (∼20x) compared to this study (∼6,300x), as our depth simulation suggests at least 4,000x is required for robust detection of low-frequency mutation in the blood.

Taken altogether, we propose a method for screening AA exposure using non-invasive substrates for early risk detection, supported by DS. Given mutations in the blood and urine are expected to be at low frequency, WGS does not have sufficient sensitivity and is therefore not recommended; likewise, low-depth duplex sequencing (for example the ∼20x used in the independent dataset) recovers SBS22a in only a minority of exposed individuals, so ultra-deep duplex sequencing is required for reliable detection in blood and urine. Duplex depth simulation analysis suggests that depths greater than 4,000x and 8,000x, for blood and urine samples, respectively, are required to reliably capture most SBS22a-positive samples. Future work in a larger, well-characterized cohort is needed to validate and support these findings. Additional work should focus on the potential dose-signature effects as well as interrogating other potential carcinogens. Overall, these findings not only expand our current understanding of AA-induced mutagenesis beyond tumors, but provide a framework for leveraging mutational processes for screening and early detection in human health and disease.

## Methods

### Samples Collection

De-identified fresh frozen cancer tissue samples with matching adjacent normal tissue were collected from patients receiving standard-of-care operations for urothelial cancer at Chang Gung Memorial Hospital, Linkou Branch, Taoyuan, Taiwan, between 2013 and 2016. Peripheral blood and mid-stream urine were collected from several patients in 2019 and 2020, several years later after receiving standard-of-care operations (**Supplemental Table 1**). In accordance with the Helsinki Declaration of 1975, written consent for research use of clinical material and clinico-pathologic data was obtained at the time of surgery. This study was approved by the Chang Gung Memorial Hospital Institutional Review Board (CGMH-IRB-102-3642A3).

### Isolation and sorting of immune cell population

Peripheral blood mononuclear cells (PBMC) were isolated by Ficoll-Paque Plus (GE Healthcare) density gradient centrifugation. Cells were stained with the following surface markers for 30 minutes at room temperature in the dark: CD45, CD3, CD56, CD19, and CD14. Live immune cells were identified based on DAPI exclusion (DAPI^-^). T-cells were defined as CD45^+^CD3^+^CD56^-^CD19^-^CD14^-^. NK-cells were defined as CD45^+^CD3^-^CD56^+^CD19^-^CD14^-^. B-cells were defined as CD45^+^CD3^-^CD56^-^CD19^+^CD14^-^. Myeloid cells were defined as CD45^+^CD3^-^CD56^-^CD19^-^CD14^+^. Cell populations were sorted with 95%-99% purity using fluorescence-activated cell sorting (FACS) Aria-III (BD Biosciences) based on cell-surface marker expression.

### Whole genome sequencing and analysis

Standard accuracy whole genome sequencing was performed on the NovaSeq 6000 platform generating 150bp PE reads. Raw sequencing data was trimmed using Trimmomatic^52^. Trimmed reads were aligned to GRCh38 and variant calling and filtering was performed as described previously^37^. Variants within 250,000 bp from blacklisted regions^53^, centromeres and telomeres were removed. For normal tissues, we removed variants detected at chromosomal locations where loss of heterozygosity was detected in the tumor using ascatNgs^54^, as these genomic locations showed high levels of germline leakage. Additionally, to distinguish somatic mutations from inherited germline variations in normal tissues, we used the binomial test to exclude variants consistent with 50% variant allele frequency expected for germline heterozygous. Probability of being a germline variant was tested using the R-call:

binom.test(x = n.mut.reads, n = total.coverage, p = 0.5, alternative = “two.sided”) where n.mut.reads is the number of reads supporting the variant allele, total.coverage is the total number of reads covering the mutation site and p is the proportion of reads that would be expected to support the mutant allele assuming it is a heterozygous variant.

### Duplex sequencing and variant calling

Duplex sequencing was performed at an average depth of 6,300x using the targeted Human Mutagenesis Panel v1.0, spanning 20 loci distributed across 20 autosomal chromosomes, totaling 48,000 bp. This panel is strategically designed to survey genomic regions that broadly reflects the trinucleotide context distribution of the whole genome while systemically avoiding confounding hotspots and non-randomly mutated genomic regions. Variant calling was performed using the associated TwinStrand Biosciences software. For samples with very limited amounts of DNA, DS was performed using 1 ng of input DNA without capture. Additionally, we removed variants found in all samples from the same patient, with at least one sample having a VAF greater than 0.1, to minimize the influence of shared background variants.

### Mutational signature analysis

VCF files were converted to mutational catalogue for mutational signature analysis using SigProfilerMatrixGenerator v1.3.5^55^. *De novo* signature extraction and decomposition into COSMIC mutational signatures v3.6 was performed using SigProfilerExtractor v1.2.6^56^. To assess signature presence and activity, COSMIC signatures identified through decomposition were used as priors with mSigAct v3.0.1.1^15^. Signature attributions contributing less than 0.05 were set to zero and the remaining signatures were renormalized to sum to 1.

### Background correction of known artifacts

We applied *SigRescueR*^57^ to remove mutational signatures associated with single-cell whole-genome sequencing artifacts and cisplatin-associated mutagenesis in relevant samples prior to *de novo* signature extraction and signature attribution. Cisplatin-associated mutational signature, SBS31^37^, was removed from patient-specific samples who received platinum-based chemotherapy as part of their standard-of-care treatment (**Supplemental Table 1**). Posterior inference was executed using *SigRescueR*’s default parameter settings.

### Simulations of duplex depth required for SBS22a detection

To infer the minimum duplex depth required for reliable detection of the exposure-associated mutational signature while reducing sequencing requirements and cost, we simulated the number of alternative duplex consensus reads supporting each mutation across different depths, ranging from 30x to 15,000x. For each simulated depth, alternative duplex consensus reads were simulated for patient-specific samples using the binomial distribution, using the simulated depth as the size and the patient-specific mutation variant allele frequency for that sample as the probability of observing the read. Mutations with 0 simulated reads were excluded from downstream analysis. We simulated each sample 10 times at each simulated depth.

### Modelling SBS22a accumulation across immune cell population

To evaluate immune cell-specific SBS22a accumulation, we fitted a negative binomial generalized linear model (GLM) with a log-link function. Immune cell type and sequencing method were included as fixed-effect predictors. To account for differences in overall mutation burden among samples, the natural logarithm of the total mutation count was included as an offset, allowing the model to estimate differences in the rate of SBS22a accumulation relative to each sample’s total mutational burden. Multiple cell-type samples were obtained from the same patient, resulting in correlated observation. To account for within-patient dependence, standard errors, confidence intervals, and p-values were estimated using Huber–White cluster-robust sandwich estimators with patient ID specified as the clustering variable, thereby reducing the risk of pseudoreplication. Myeloid samples were excluded from the GLM analysis due to absence in SBS22a mutations, resulting in an unestimable boundary effect that would destabilize parameter estimation. The final model was specified as resulting in:

SBS22a_counts ∼ immune_cell_type_NK-cell_ + sequencing_method + offset[ln(total_mutations)]

using a negative binomial variance parameter of LJ=1 and patient-clustered robust standard errors. Statistical analyses were performed in Python using pandas for data processing, NumPy for numerical calculations, and statsmodels for model fitting and statistical inference. Regression coefficients and 95% confidence intervals were visualized as a forest plot using ggplot2.

### Statistical analysis

For all statistical analyses involving mutational signatures, signature presence was defined as a signature accounting for more than 5% of the total mutational contribution within a sample. To assess whether pairs of mutational signatures co-occurred more frequently than due to chance, pairwise co-occurrence was performed based on the presence or absence of the signature across all samples using two-sided Fisher’s exact test. Sample-specific signature enrichment was assessed based on the presence or absence of the signature between groups using two-sided Fisher’s exact test. The odds ratio was computed using Haldane-Anscombe correction to account for zero cell counts. A one-sided exact binomial test was used to assess whether a signature was present in more than 50% of the samples (null hypothesis). Spearman’s rank correlation was used to assess the association of clonal and nonclonal SBS22a and aging-associated signature with the expected gradient of cellular heterogeneity, increasing from tumor to normal, blood and urine. A two-sided binomial test was used to evaluate differences in SBS22a attribution for clonal and nonclonal mutation in patient-specific samples.

## Data Availability

All data produced in the present study are available upon reasonable request to the authors

https://github.com/ZhivaguiLab/AAmanu

https://ega-archive.org/datasets/EGAD00001003467

## Data Availability

Bulk and single-cell WGS and duplex sequencing data that support the findings of this study have been deposited in EGAC00001000708. The data underlying the figure and in-text analyses are provided in the Supplemental Tables. All other data are available on request from the corresponding author.

## Code Availability

All code used to generate the figures and analyses reported in this manuscript is openly available on GitHub at: https://github.com/ZhivaguiLab/AAmanu. Detailed instructions for reproducing the results are provided in the repository’s README file.

## Funding

Singapore National Medical Research Council grants NMRC/CIRG/1422/2015 and MOH-000032/MOH-CIRG18may-0004, the Singapore Ministry of Health via the Duke-NUS Signature Research Programmes (SGR), institutional start-up funds from the University of Nevada Las Vegas, and Centers of Biomedical Research Excellence (COBRE) Phase I grant P20 GM121325 from the National Institutes of Health.

## Acknowledgements

The illustrations were created by BioRender.com.

## Author Contributions

A.B., W.Y., M.L. and N.J. conducted the experiments. P.N., A.B., S.S. conducted the analysis. P.L. and J.P. collected the tissue, blood and urine samples. E.S., C.V., J.S., and

F.L. performed sequencing. V.C. performed FACS of PBMC. P.N., M.Z., S.R., A.B., and S.S. drafted and reviewed the final manuscript. All authors approved the final version of the manuscript.

## Competing Interests

E.S., C.V., J.S., and F.L. were employees of and equity holders in TwinStrand Biosciences, Inc. at the time this work was done. The other authors declare that they have no competing interests.

---

**Supplemental Figure 1. Distribution of variant allele frequency across tumor and normal tissues.** Histogram illustrating the variant allele frequency by tissue type per individual, where the x-axis represents the binned allele frequency and the y-axis represents the number of variants within each binned. Bars are colored by sample type.

**Supplemental Figure 2. Transcription strand bias analysis for whole-genome sequenced tumor and normal samples per individual.** Mutational profiles were stratified by individual and sample type. Bars (left y-axis) shows the proportion of mutations occurring on the transcribed (T) and non-transcribed (N) strand, with color bar representing the six SBS mutation contexts. The dashed line (right y-axis) represents the p-value from the strand bias analysis (two-sided binomial test; null hypothesis: p=0.5), with significance levels indicated by asterisks (*, p<0.05; **, p<0.01; ***, p<0.001; ****, p<0.0001).

**Supplemental Figure 3. Co-occurrence analysis across COSMIC signature.** Fisher’s exact test was used to assess whether the detection of one signature is associated with the detection of another signature. The color gradient represents the log odds ratio, with blue indicating depletion of co-occurrence (OR < 0), white indicating no association (OR = 0), and red indicating enrichment of co-occurrence (OR > 0).

**Supplemental Figure 4. Transcription strand bias analysis for duplex sequenced tumor and normal samples per individual.** Mutational profiles were stratified by individual and sample type. Bars (left y-axis) shows the proportion of mutations occurring on the transcribed (T) and non-transcribed (N) strand, with color bar representing the six SBS mutation contexts. The dashed line (right y-axis) represents the p-value from the strand bias analysis (two-sided binomial test; null hypothesis: p=0.5), with significance levels indicated by asterisks (*, p<0.05; **, p<0.01; ***, p<0.001; ****, p<0.0001).

**Supplemental Figure 5. Somatic mutation frequencies of duplex sequenced samples at the SBS, DBS, and ID level.** Y-axis shows log mutation per megabase. Each point represents an individual sample, ranked lowest to highest by mutation count, and colored by sample type. Horizontal line represents the median mutational burden for each mutation type.

**Supplemental Figure 6. Transcription strand bias analysis for duplex sequenced blood and urine samples per individual.** Mutational profiles were stratified by individual and sample type. Bars (left y-axis) shows the proportion of mutations occurring on the transcribed (T) and non-transcribed (N) strand, with color bar representing the six SBS mutation contexts. The dashed line (right y-axis) represents the p-value from the strand bias analysis (two-sided binomial test; null hypothesis: p=0.5), with significance levels indicated by asterisks (*, p<0.05; **, p<0.01).

**Supplemental Figure 7. Probability of observed read count across clone fractions.** The probability of observing greater than or equal to the detected read count at each simulated cell population was computed using the hypergeometric distribution. The number of successes (m) and failures (n) in the population were defined as the number of cells with read counts greater than or equal to the detected read count and below detected read count, respectively, based on 100ng of input DNA. The number of draws (k) was selected using the median duplex coverage. X-axis represents the proportion of cells carrying the mutation, ranging from 0.000001 to 0.005. Y-axis represents the probability of observing greater than or equal to the detected read count. Trace plot illustrates how increasing the clone fraction increases the probability of detecting duplex supporting reads at each threshold.

**Supplemental Figure 8. Overlap of mutations across patient-derived samples with blood-derived variants.** Venn diagram illustrating the number of mutations shared across samples for the same patient.

**Supplemental Figure 9. Background correction of immune sorted fraction whole-genome sequenced.** (**A**) SBS-96 mutational signature plot showing the uncleaned (Original), control (Background), background corrected spectra (Cleaned), and reconstructed profile from the T-cell of UC18. Alpha transparency indicates background (a = 0.5) and background corrected spectra (a = 1) levels in the reconstructed profile. (**B**) Scatter plot summarizing relative background activity estimated. Each point represents a single cell, with shape denoting the cell type and color indicating the patient.

**Supplemental Figure 10. Mutational spectra for single-cell whole-genome sequenced immune cells per individual after background correction.** Mutational profiles were stratified by individual and sample type. SBS96 mutational profile, with color bar representing the six SBS mutation contexts.

**Supplemental Figure 11. Estimated SBS31 in duplex sequenced immune cell populations of UC14.** SBS96 mutational profile, with color bar representing the six SBS mutation contexts. Mutational profiles were annotated using varying alpha transparency, where alpha 0.5 and 1 indicate SBS31 and non-SBS31 mutations, respectively. The estimated SBS31 contribution is displayed within the plot.

**Supplemental Figure 12. Mutational spectra for duplex sequenced immune cell populations per individual.** Mutational profiles were stratified by individual and sample type. SBS96 mutational profile, with color bar representing the six SBS mutation contexts.

**Supplemental Figure 13. Distribution of clonal mutation across PBMC and sorted immune cell population.** Barplot showing the distribution of clonal (yellow) and nonclonal mutations (grey) stratified by PBMC-exclusive mutations (“No”) and those found in both PBMC and sorted immune fraction (“Yes”) for UC7 and UC14.

## Notes

### Author Declarations

This study was approved by the Chang Gung Memorial Hospital Institutional Review Board (CGMH-IRB-102-3642A3).

## References

1. Alexandrov, L. B. et al. Signatures of mutational processes in human cancer. Nature 500, 415–421 (2013).

2. Alexandrov, L. B. et al. The repertoire of mutational signatures in human cancer. Nature 578, 94–101 (2020).

3. Martincorena, I. Somatic mutation and clonal expansions in human tissues. Genome Med. 11, 35 (2019).

4. Moore, L. et al. The mutational landscape of human somatic and germline cells. Nature 597, 381–386 (2021).

5. Díaz-Gay, M. et al. Geographic and age variations in mutational processes in colorectal cancer. Nature 643, 230–240 (2025).

6. Lu, H. et al. Aristolochic acid mutational signature defines the low-risk subtype in upper tract urothelial carcinoma. Theranostics 10, 4323–4333 (2020).

7. Schiffman, J. D., Fisher, P. G. & Gibbs, P. Early detection of cancer: past, present, and future. Am. Soc. Clin. Oncol. Educ. Book 35, 57–65 (2015).

8. Debelle, F. D., Vanherweghem, J.-L. & Nortier, J. L. Aristolochic acid nephropathy: a worldwide problem. Kidney Int. 74, 158–169 (2008).

9. Michl, J., Ingrouille, M. J., Simmonds, M. S. J. & Heinrich, M. Naturally occurring aristolochic acid analogues and their toxicities. Nat. Prod. Rep. 31, 676–693 (2014).

10. Michl, J. et al. LC-MS- and (1)H NMR-based metabolomic analysis and in vitro toxicological assessment of 43 Aristolochia species. J. Nat. Prod. 79, 30–37 (2016).

11. Das, S. et al. Aristolochic acid-associated cancers: a public health risk in need of global action. Nat. Rev. Cancer 22, 576–591 (2022).

12. Hoang, M. L. et al. Mutational signature of aristolochic acid exposure as revealed by whole-exome sequencing. Sci. Transl. Med. 5, 197ra102 (2013).

13. Poon, S. L. et al. Genome-wide mutational signatures of aristolochic acid and its application as a screening tool. Sci. Transl. Med. 5, 197ra101 (2013).

14. Poon, S. L. et al. Mutation signatures implicate aristolochic acid in bladder cancer development. Genome Med. 7, 38 (2015).

15. Ng, A. W. T. et al. Aristolochic acids and their derivatives are widely implicated in liver cancers in Taiwan and throughout Asia. Sci. Transl. Med. 9, eaan6446 (2017).

16. Chen, L. et al. Deep whole-genome analysis of 494 hepatocellular carcinomas. Nature 627, 586–593 (2024).

17. Brunner, S. F. et al. Somatic mutations and clonal dynamics in healthy and cirrhotic human liver. Nature 574, 538–542 (2019).

18. Li, R. et al. Macroscopic somatic clonal expansion in morphologically normal human urothelium. Science 370, 82–89 (2020).

19. Salk, J. J., Schmitt, M. W. & Loeb, L. A. Enhancing the accuracy of next-generation sequencing for detecting rare and subclonal mutations. Nat. Rev. Genet. 19, 269–285 (2018).

20. Satyal, U., Srivastava, A. & Abbosh, P. H. Urine biopsy-liquid gold for molecular detection and surveillance of bladder cancer. Front. Oncol. 9, 1266 (2019).

21. Menon, V. & Brash, D. E. Next-generation sequencing methodologies to detect low-frequency mutations: ‘Catch me if you can’. Mutat. Res. Rev. Mutat. Res. 792, 108471 (2023).

22. Salk, J. J. & Kennedy, S. R. Next-generation genotoxicology: Using modern sequencing technologies to assess somatic Mutagenesis and cancer risk. Environ. Mol. Mutagen. 61, 135–151 (2020).

23. Wang, Y., et al. Systemic mutagen exposures reported by normal kidney cell genomes. bioRxiv (2026) doi:10.64898/2026.04.07.716715.

24. Senkin, S. et al. Geographic variation of mutagenic exposures in kidney cancer genomes. Nature 629, 910–918 (2024).

25. Nik-Zainal, S. et al. Mutational processes molding the genomes of 21 breast cancers. Cell 149, 979–993 (2012).

26. Wang, Y. et al. Genetic toxicity testing using human in vitro organotypic airway cultures: Assessing DNA damage with the CometChip and mutagenesis by Duplex Sequencing. Environ. Mol. Mutagen. 62, 306–318 (2021).

27. Hiley, C., de Bruin, E. C., McGranahan, N. & Swanton, C. Deciphering intratumor heterogeneity and temporal acquisition of driver events to refine precision medicine. Genome Biol. 15, 453 (2014).

28. Tomasetti, C. & Vogelstein, B. Cancer etiology. Variation in cancer risk among tissues can be explained by the number of stem cell divisions. Science 347, 78–81 (2015).

29. Ding, Y. C., Tao, S., Mao, A., Ziv, E. & Neuhausen, S. Cosmic signature SBS39 is associated with homologous recombination deficiency. medRxiv (2024) doi:10.1101/2024.10.23.24316019.

30. Degasperi, A. et al. Substitution mutational signatures in whole-genome-sequenced cancers in the UK population. Science 376, science.abl9283 (2022).

31. Díaz-Gay, M. et al. The mutagenic forces shaping the genomes of lung cancer in never smokers. Nature 644, 133–144 (2025).

32. Zhang, T., et al. Deciphering lung adenocarcinoma evolution and the role of LINE-1 retrotransposition. bioRxiv(2025). doi: 10.1101/2025.03.14.643063.

33. Costello, M. et al. Discovery and characterization of artifactual mutations in deep coverage targeted capture sequencing data due to oxidative DNA damage during sample preparation. Nucleic Acids Res. 41, e67 (2013).

34. Chen, C. K., Liao, J., Li, M. S. & Khoo, B. L. Urine biopsy technologies: Cancer and beyond. Theranostics 10, 7872–7888 (2020).

35. Oshi, M. et al. Urine as a source of liquid biopsy for cancer. Cancers (Basel) 13, 2652 (2021).

36. Lawson, A. R. J. et al. Extensive heterogeneity in somatic mutation and selection in the human bladder. Science 370, 75–82 (2020).

37. Boot, A. et al. In-depth characterization of the cisplatin mutational signature in human cell lines and in esophageal and liver tumors. Genome Res. 28, 654–665 (2018).

38. Springer, S. U. et al. Non-invasive detection of urothelial cancer through the analysis of driver gene mutations and aneuploidy. Elife 7, e32143 (2018).

39. Kennedy, S. R. et al. Detecting ultralow-frequency mutations by Duplex Sequencing. Nat. Protoc. 9, 2586–2606 (2014).

40. Behjati, S. et al. Genome sequencing of normal cells reveals developmental lineages and mutational processes. Nature 513, 422–425 (2014).

41. Lee-Six, H. et al. The landscape of somatic mutation in normal colorectal epithelial cells. Nature 574, 532–537 (2019).

42. Fabre, M. A. et al. The longitudinal dynamics and natural history of clonal haematopoiesis. Nature 606, 335–342 (2022).

43. Liu, Y., Chan, C.-K., Jin, L., Wong, S.-K. & Chan, W. Quantitation of DNA adducts in target and nontarget organs of aristolochic acid I-exposed rats: Correlating DNA adduct levels with organotropic activities. Chem. Res. Toxicol. 32, 397–399 (2019).

44. Ham, Y.-H. et al. Positive feedback mechanism in aristolochic acid I exposure-induced anemia and DNA adduct formation: Implications for Balkan endemic nephropathy. J. Agric. Food Chem. 72, 18155–18161 (2024).

45. Jelaković, B. et al. Aristolactam-DNA adducts are a biomarker of environmental exposure to aristolochic acid. Kidney Int. 81, 559–567 (2012).

46. Sidorenko, V. S. et al. Lack of recognition by global-genome nucleotide excision repair accounts for the high mutagenicity and persistence of aristolactam-DNA adducts. Nucleic Acids Res. 40, 2494–2505 (2012).

47. Schmeiser, H. H. et al. Exceptionally long-term persistence of DNA adducts formed by carcinogenic aristolochic acid I in renal tissue from patients with aristolochic acid nephropathy. Int. J. Cancer 135, 502–507 (2014).

48. Zhang, L. et al. Single-cell whole-genome sequencing reveals the functional landscape of somatic mutations in B lymphocytes across the human lifespan. Proc. Natl. Acad. Sci. U. S. A. 116, 9014–9019 (2019).

49. Machado, H. E. et al. Diverse mutational landscapes in human lymphocytes. Nature 608, 724–732 (2022).

50. Sprent, J. Lifespans of naive, memory and effector lymphocytes. Curr. Opin. Immunol. 5, 433–438 (1993).

51. He, Z. et al. Rapid turnover and high production rate of myeloid cells in adult rhesus macaques with compensations during aging. J. Immunol. 200, 4059–4067 (2018).

52. Bolger, A. M., Lohse, M. & Usadel, B. Trimmomatic: a flexible trimmer for Illumina sequence data. Bioinformatics 30, 2114–2120 (2014).

53. Amemiya, H. M., Kundaje, A. & Boyle, A. P. The ENCODE blacklist: Identification of problematic regions of the genome. Sci. Rep. 9, 9354 (2019).

54. Raine, K. M. et al. AscatNgs: Identifying somatically acquired copy-number alterations from whole-genome sequencing data: AscatNgs: Identifying copy-number alterations from sequencing data. Curr. Protoc. Bioinformatics 56, 15.9.1–15.9.17 (2016).

55. Bergstrom, E. N. et al. SigProfilerMatrixGenerator: a tool for visualizing and exploring patterns of small mutational events. BMC Genomics 20, 685 (2019).

56. Islam, S. M. A. et al. Uncovering novel mutational signatures by de novo extraction with SigProfilerExtractor. Cell Genom. 2, None (2022).

57. Nguyen, P. T. & Zhivagui, M. SigRescueR: a pan-system framework for noise correction and mutational signature identification across sequencing platforms. Brief. Bioinform. 27, bbag099 (2026).

