## Supplementary figures and images for "Non-invasive detection of mutagenic exposure to aristolochic acid using ultra-deep sequencing of blood and urine samples"

### Suppl. Fig1

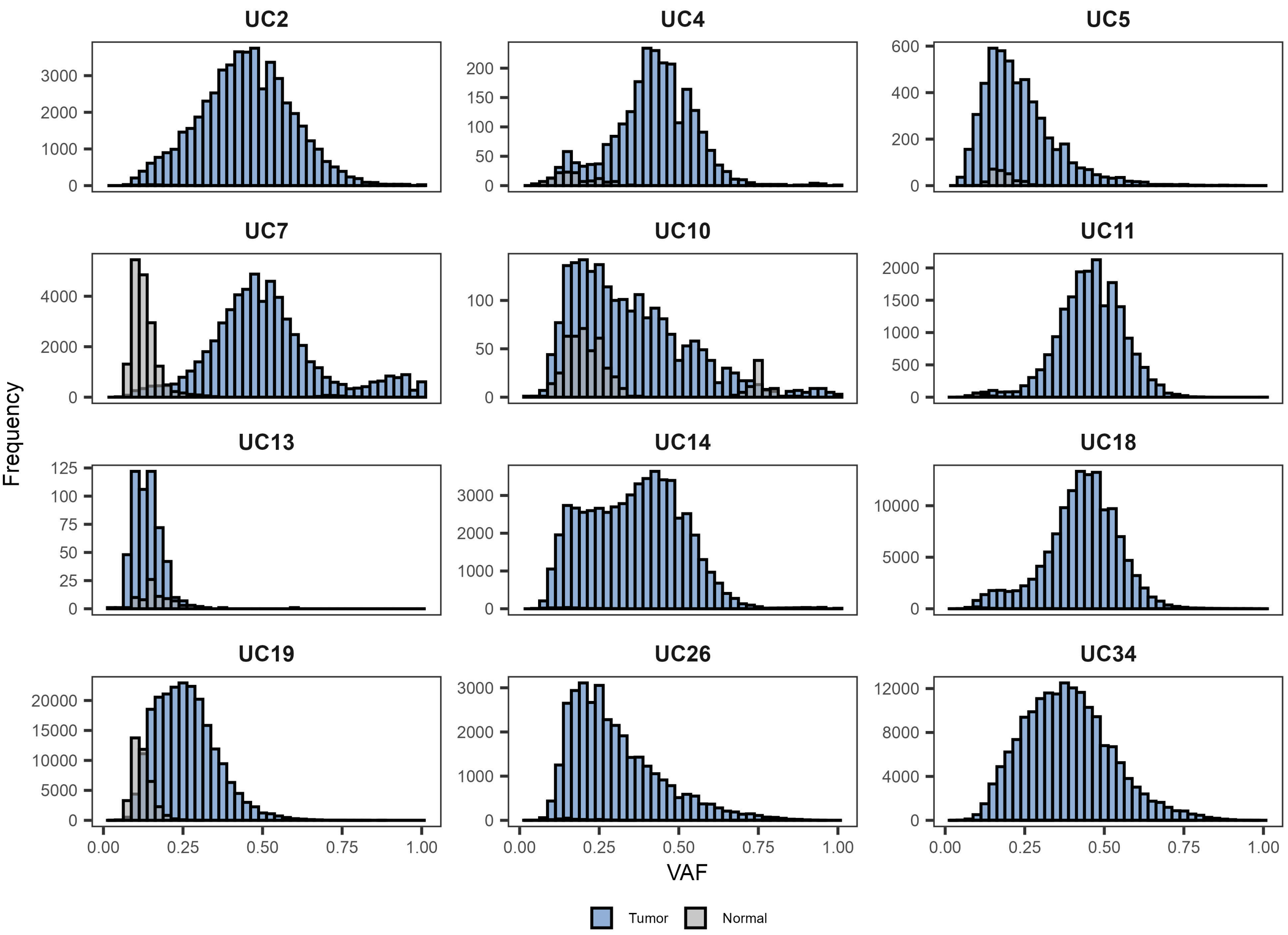

### Suppl. Fig2

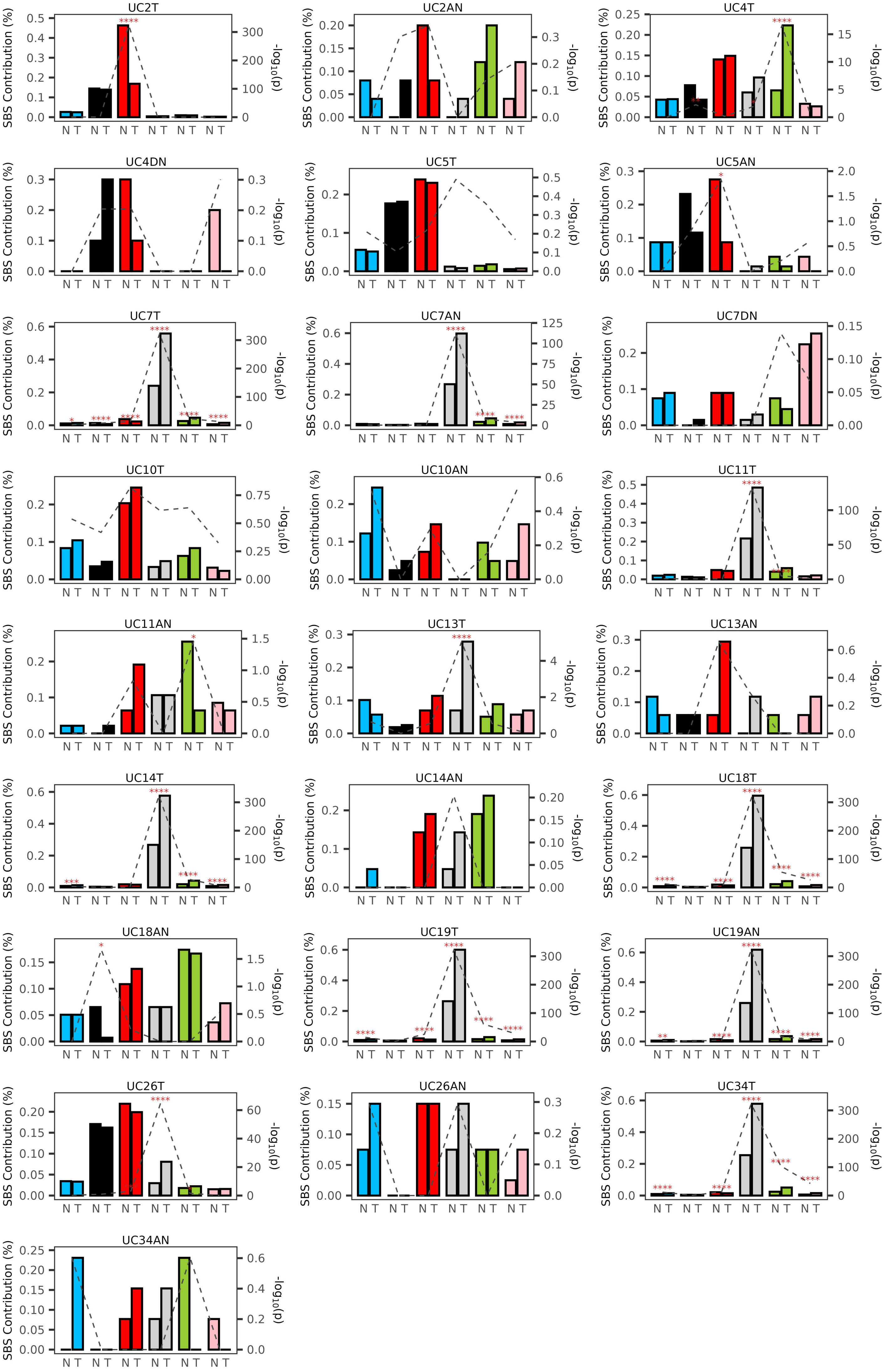

### Suppl. Fig3

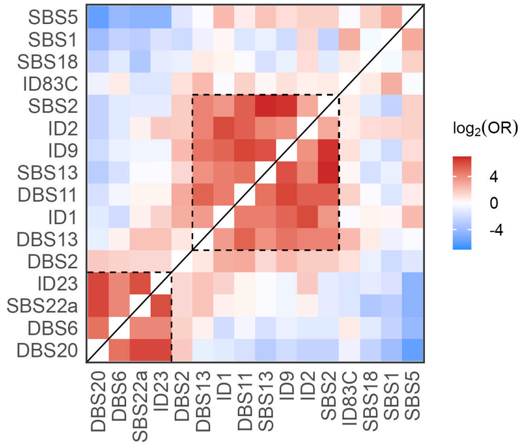

### Suppl. Fig4

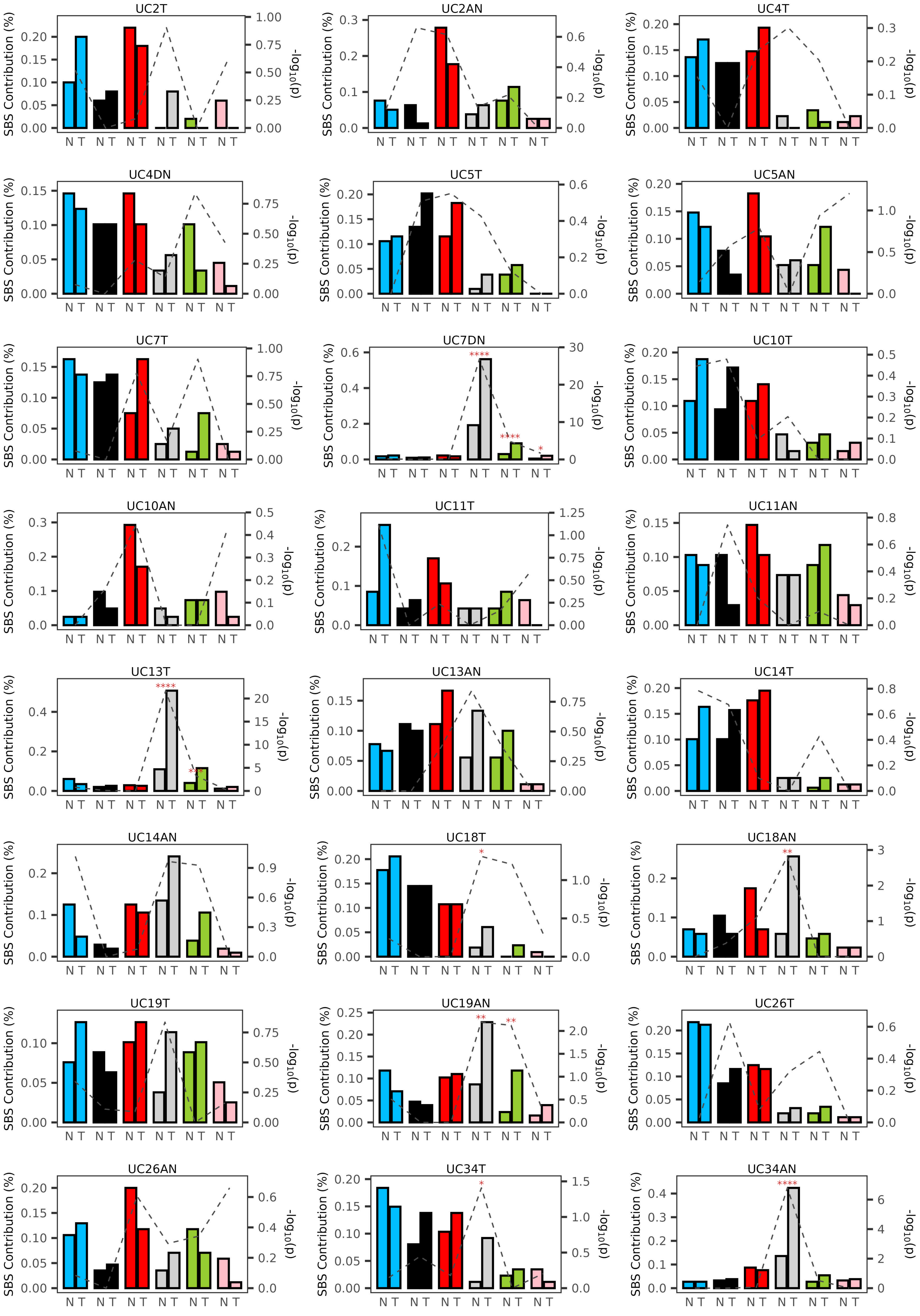

### Suppl. Fig5

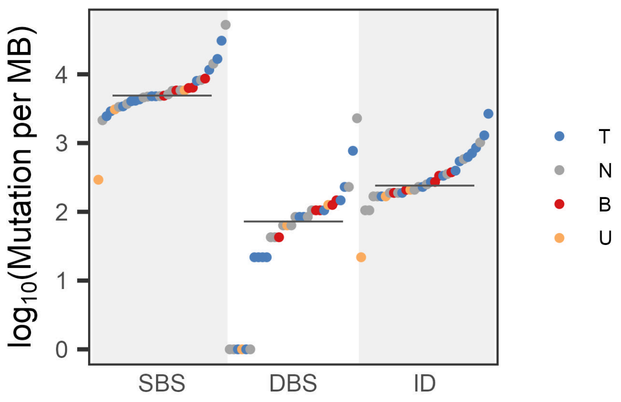

### Suppl. Fig6

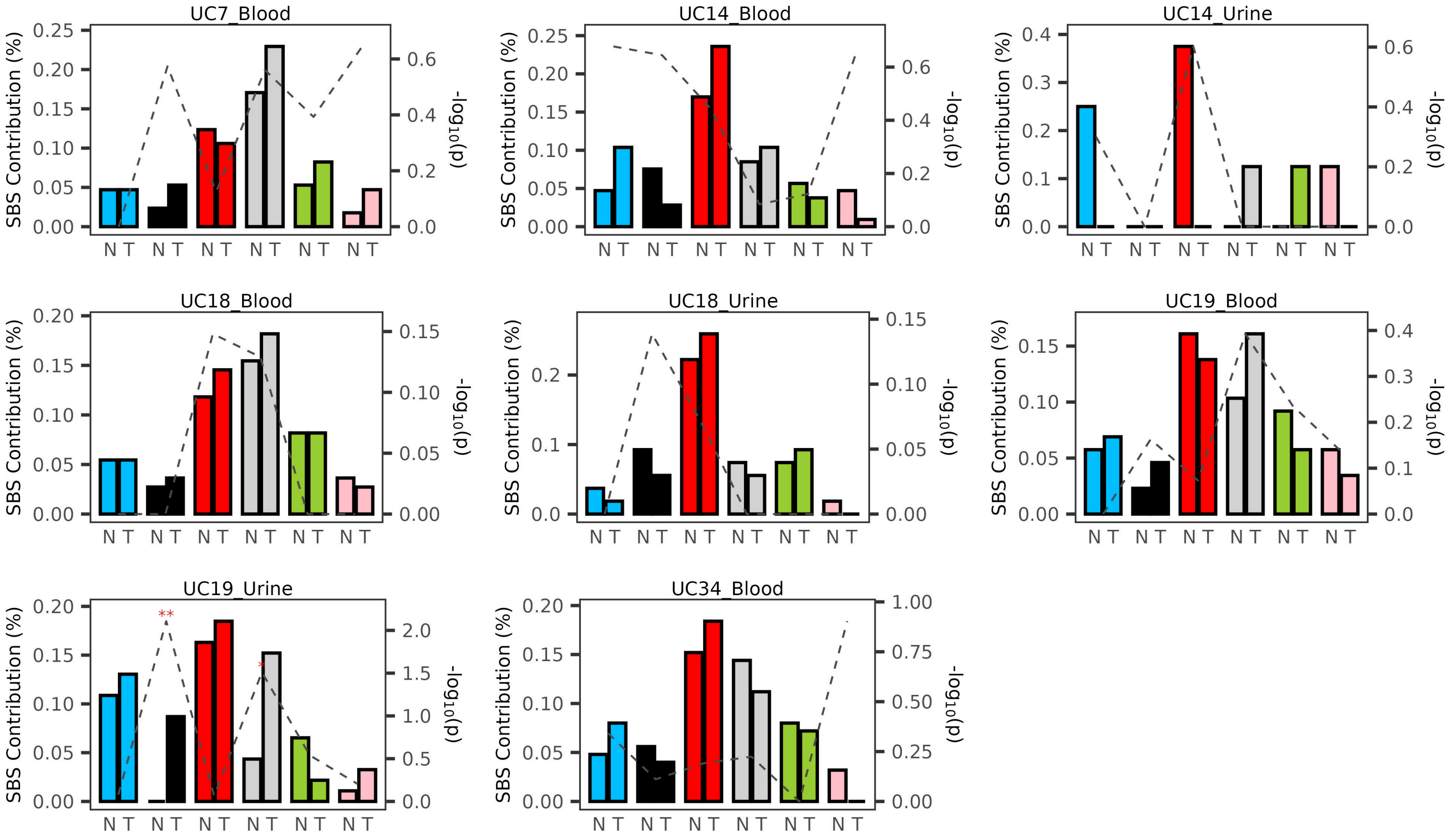

### Suppl. Fig7

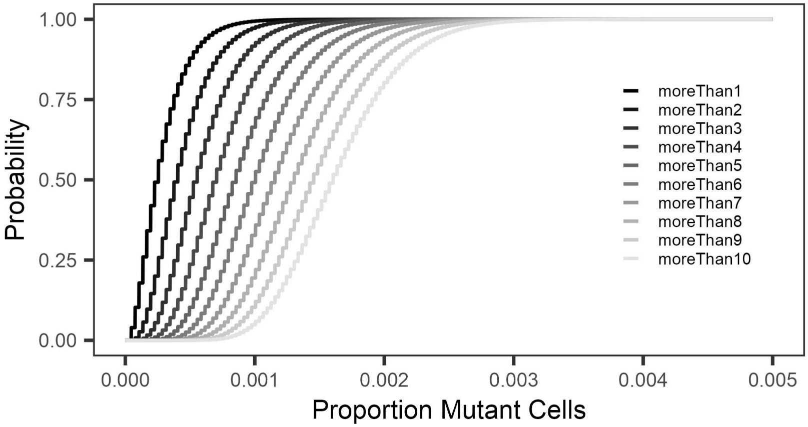

### Suppl. Fig8

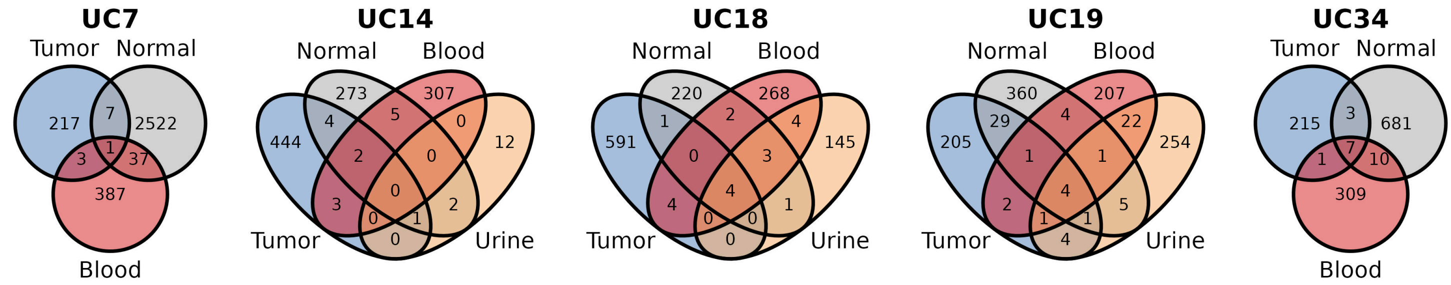

### Suppl. Fig9

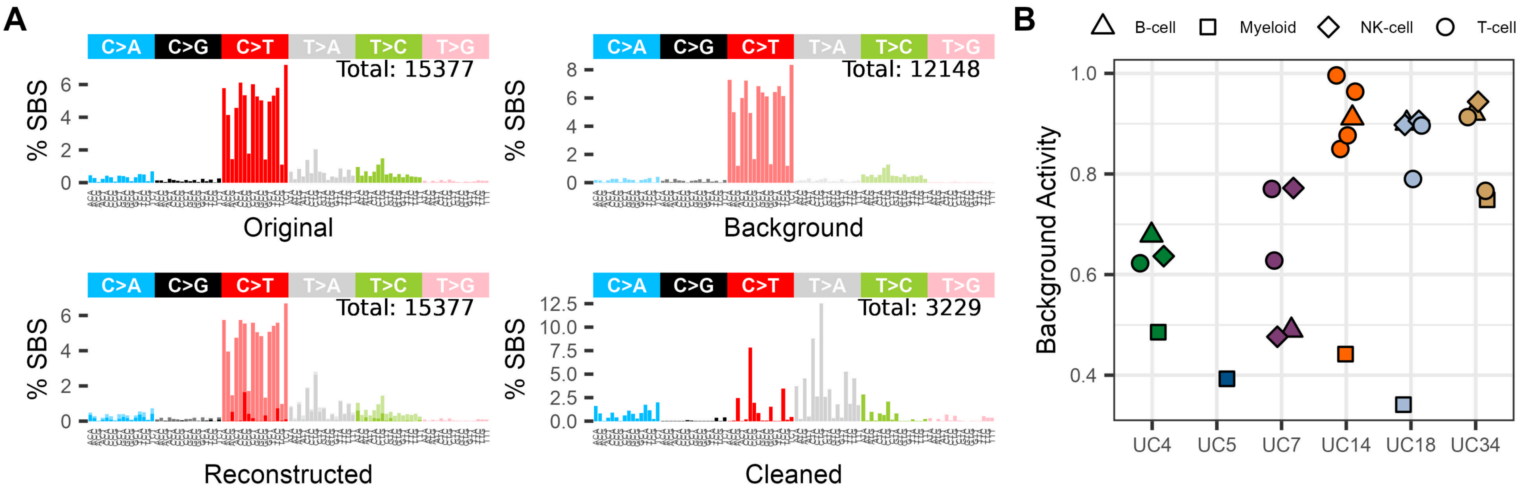

### Suppl. Fig10

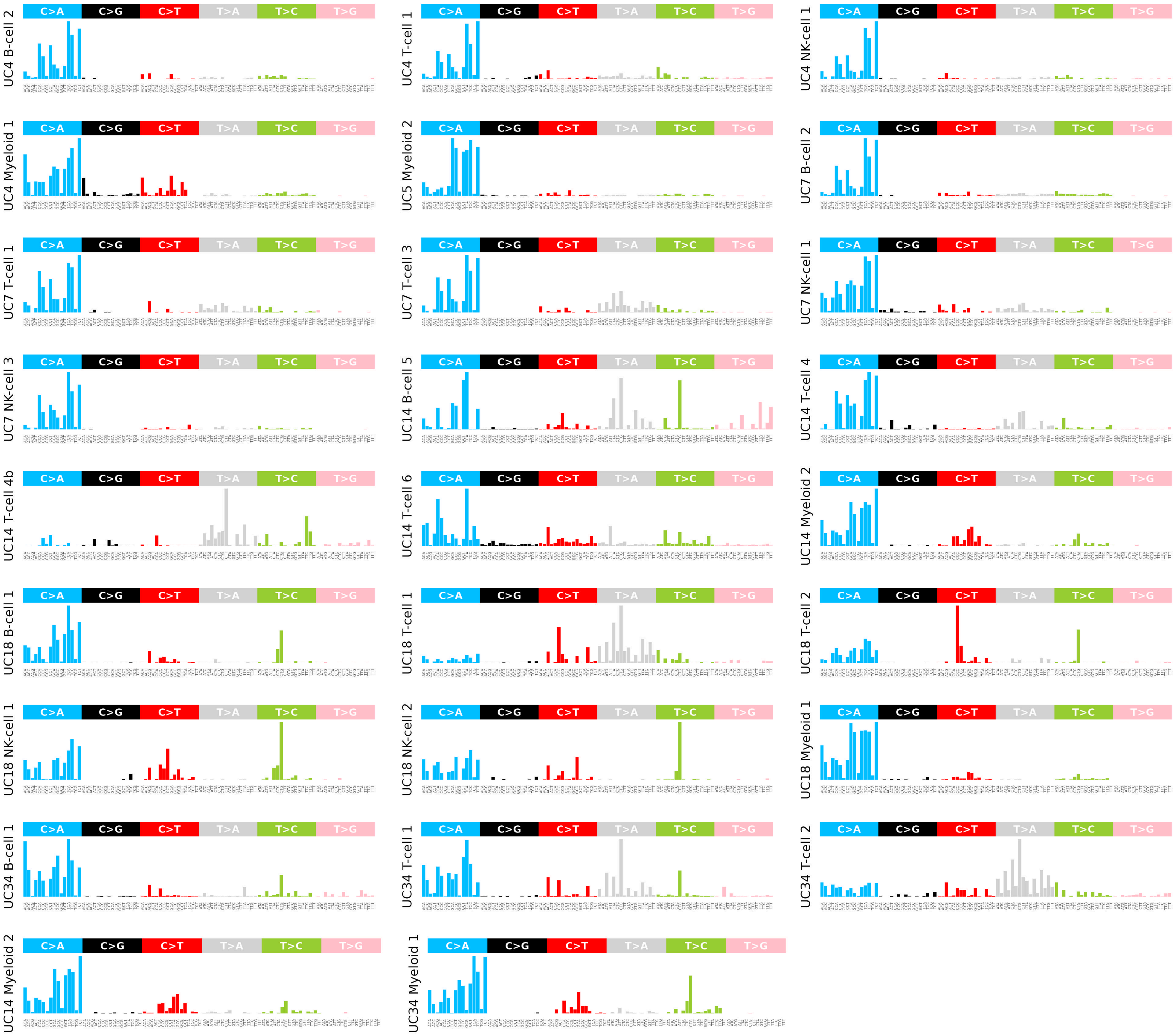

### Suppl. Fig11

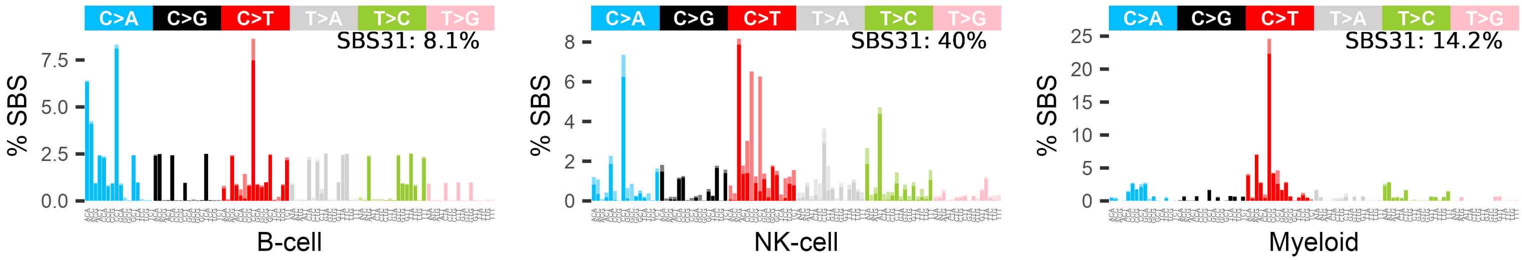

### Suppl. Fig12

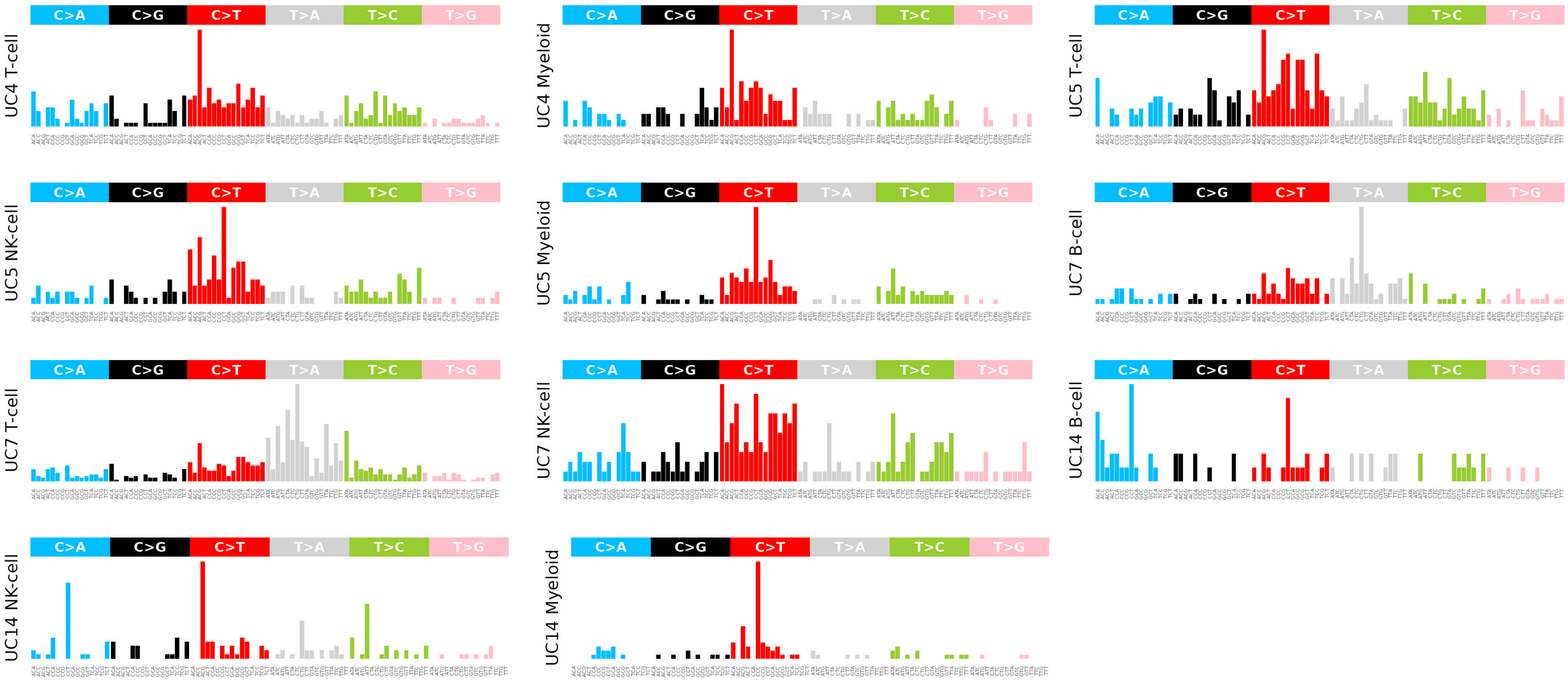

### Suppl. Fig13

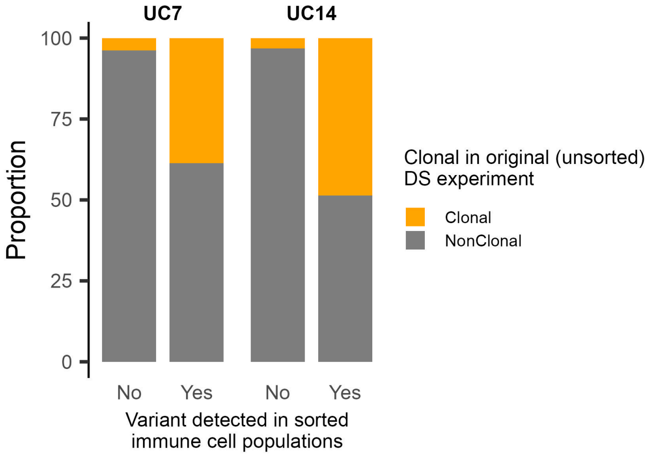
